# Empirical validation of a predicted emotional modulation dimension linking temporal variability and individual differences in PTSD

**DOI:** 10.64898/2026.08.05.26359316

**Authors:** Toshinori Chiba, Masaya Ito, Masaya Ichii, Kentarou Ide, Misa Murakami, Takero Terayama, Takatomi Kubo, Keiichiro Nishida, Nao Kobayashi, Taku Saito, Yuriko Takagishi, Florentine H. S. van der Does, Hironori Kuga, Masaru Horikoshi, Miyako Shirakawa-Nishi, Taishiro Kishimoto, Hiroyuki Toda, Tetsufumi Kanazawa, Nic J. van der Wee, Noam Goldway, Aurelio Cortese, Erik J. Giltay, Masanori Nagamine, Petra Ritter, Eric Vermetten, Talma Hendler, Mitsuo Kawato

## Abstract

Current dimensional approaches to psychiatric disorders have largely focused on explaining differences between individuals, whereas it remains unknown whether symptom dynamics within individuals are organized by the same underlying dimensions.

In PTSD, temporal symptom variability may represent a clinically meaningful source of heterogeneity relevant to spontaneous recovery, chronicity, and treatment response. Our reciprocal inhibition model of PTSD proposed that both between-individual heterogeneity and within-individual dynamics may be organized along a dimension reflecting the relative balance between re-experiencing and avoidance symptoms (symptom imbalance), potentially corresponding to shifts between states of emotional under- and overmodulation.

Here, using seven longitudinal and two cross-sectional PTSD cohorts spanning disorder development, chronicity, and recovery, we examined whether symptom heterogeneity between individuals and within individuals over time is organized along shared latent symptom dimensions. Principal component analysis (PCA) performed separately on between-individual variability (individual differences) and within-individual variation (temporal variability) consistently recovered the same two axes: the first indexing overall symptom severity and the second reflecting the proposed symptom imbalance.

To enable direct comparison across cohorts and between-individual and temporal scales, we integrated cohort-specific covariance structures using hierarchical multi-group PCA yielding universal axes (uPC1/uPC2). Mapping treatment trajectories onto this shared symptom space revealed that two first-line psychotherapies—cognitive processing therapy (CPT) and eye movement desensitization and reprocessing (EMDR)—produced comparable reductions in overall symptom severity (uPC1), but opposite shifts along symptom imbalance (uPC2). These findings suggest treatment-related symptom trajectories that are not captured by severity alone and provide a quantitative basis for treatment stratification grounded in symptom imbalance dynamics, motivating prospective tests of state-dependent intervention in PTSD and related psychiatric disorders.

## INTRODUCTION

Psychiatric disorders exhibit substantial between-individual heterogeneity, and diagnostic boundaries are often indistinct. Even within a single diagnosis, patients can present with highly diverse symptom profiles, reflecting the combinatorial complexity of current diagnostic systems^1^. Such heterogeneity is clinically important because patients with similar diagnoses and overall disorder related severity, often respond differently to the same intervention, highlighting the limitations of severity-based models of psychopathology and treatment management^2,3^. This clinically manifested complexity have motivated dimensional frameworks such as the Research Domain Criteria (RDoC) and the Hierarchical Taxonomy of Psychopathology (HiTOP), which propose that psychopathology may be better understood in terms of dimensional structure and transdiagnostic domain abnormalities rather than discrete diagnostic categories and symptom severity^4,5^. Inspired by these frameworks, several studies including in depression and PTSD have reported symptom- or circuit-based biotypes associated with differential treatment outcomes, suggesting that heterogeneity may carry clinically actionable information beyond overall symptom severity^6–11^.

Existing dimensional approaches have focused primarily on between-individual differences and do not account for temporal dynamics within individuals. Notably, the reproducibility of several proposed symptom-based and neurobiological subtypes has proven challenging^12,13^, raising the possibility that clinically relevant heterogeneity may not be fully captured by stable between-individual differences alone. Accumulating studies have shown that psychiatric symptoms exhibit substantial within-individual dynamics^14–18^. Recent network- and trajectory-based approaches have shown that within-individual symptom dynamics are structured rather than random and may partially overlap with between-individual symptom network structure^18–20^. At the same time, these and related studies have also highlighted important discrepancies between within-individual and between-individual network structures^21^, leaving unresolved whether within-individual dynamics and between-individual heterogeneity share common latent symptom dimensions. Establishing such latent symptom dimensions would allow knowledge accumulated from between-individual heterogeneity to inform within-individual dynamics and facilitate identification of underlying biological mechanisms that could inform process-based interventions. If an individual’s state/position shifts along treatment-relevant dimensions over time, treatment efficacy itself may vary accordingly.

PTSD provides a useful model to address this question because it is characterized by substantial heterogeneity in symptom expression, neurobiology, and treatment response^22,23^, while theoretical frameworks have previously suggested that clinically meaningful symptom variation may be organized along a low-dimensional manifold independent of overall symptom severity. Building on models proposing that between-individual heterogeneity in PTSD symptom presentations is organized along a continuum from emotional under- to overmodulation^24,25^, we previously hypothesized that this continuum may be observable at the symptom level. Specifically, we proposed “symptom imbalance” as the relative balance between re-experiencing and avoidance symptom clusters, hypothesized to reflect emotional under- and overmodulation, respectively^26^. Within this framework, we proposed a dynamical model in which transitions along this under- and over-modulation continuum could account for within-individual clinical state dynamics that accompany temporal fluctuations in symptom organization^26^. This framework therefore predicts that symptom imbalance should emerge as a shared dimension underlying both between-individual heterogeneity and within-individual dynamics.

Here, we examine whether the previously hypothesized symptom imbalance emerges from unsupervised decomposition of symptom covariance patterns derived separately from between-individual and temporal covariance matrices. We further test whether this dimension extends beyond clinically diagnosed PTSD to non-clinical populations reporting stress-related symptoms, and whether different psychotherapy based treatment approaches are associated with distinct trajectories along this dimension. Establishing such a shared dimension may open avenues for state-dependent intervention strategies, in which treatment response may depend not only on overall symptom severity but also on an individual’s current position along this severity-independent dimension.

## RESULTS

### Dataset overview

The main analyses included nine cohorts (seven longitudinal and two cross-sectional), comprising 24,628 eligible participants for between-individual analyses and 67,055 repeated observations for within-individual analyses (see Methods, **Table 1**, **Supplementary Table 1**). The longitudinal cohorts spanned multiple phases of trauma-related symptom trajectories, including post-trauma symptom development with clearly defined trauma onset (Development phase: D-1–D-3), chronic symptom maintenance in individuals with established PTSD (Maintenance phase: M-1–M-2), and treatment-related recovery during trauma-focused psychotherapy (Recovery phase: R-1–R-2). These cohorts varied substantially in trauma type and sampling interval, with assessment intervals ranging from days to years, including one intensive longitudinal cohort (M-2, n = 23) in which symptoms were assessed every five days over a six-month period (**Table 1**). Two independent cross-sectional trauma-exposed online cohorts were included to examine whether the between-individual covariance structure generalized across larger and more diverse populations, including one predominantly non-Japanese cohort (O-1, n = 887) and one Japanese cohort (O-2, n = 4,392).

**Table 1.**
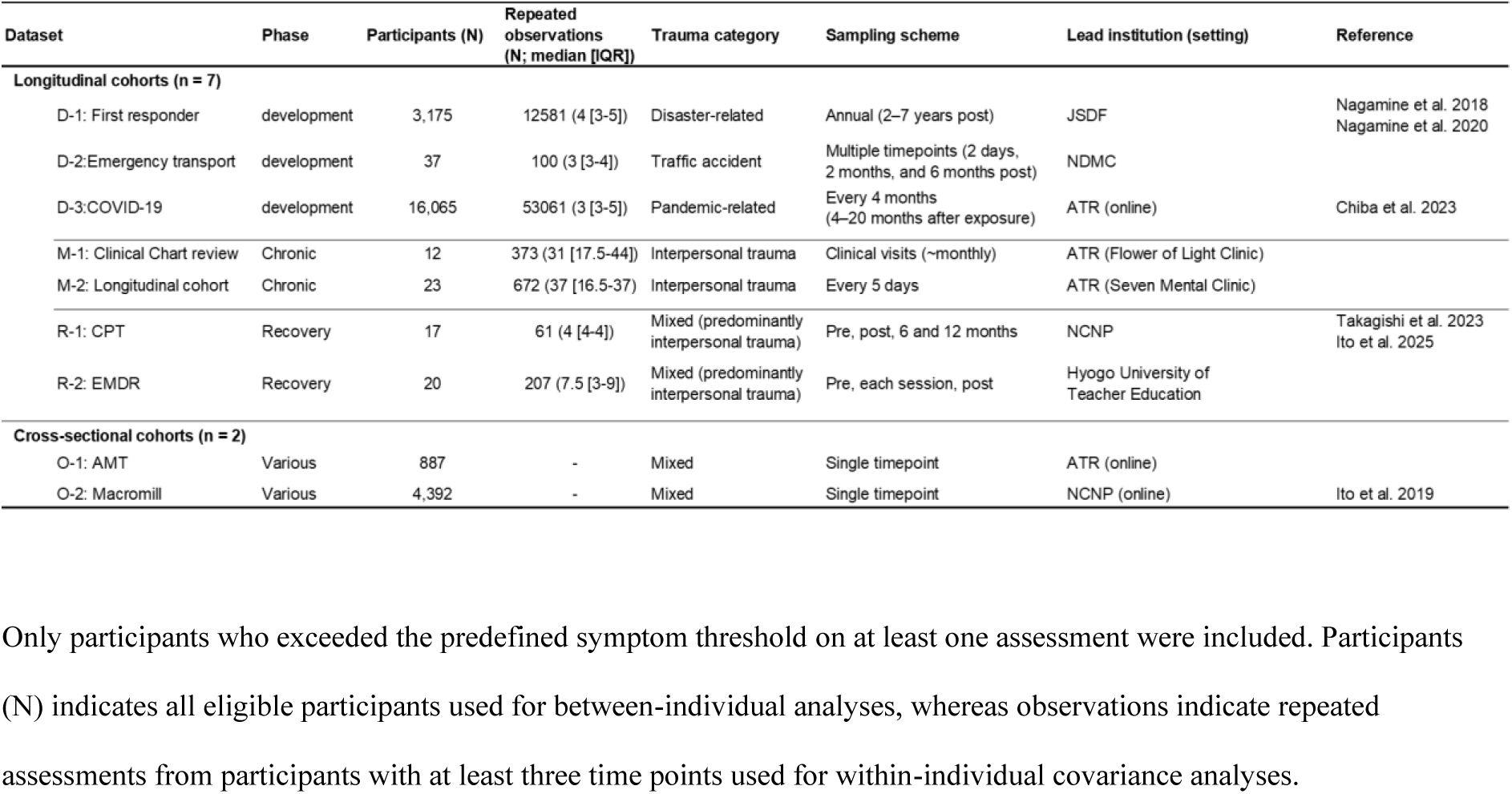
Cohorts included in the main analyses.

Despite substantial differences in cohort characteristics, clinical context, and temporal resolution, all cohorts used the same questionnaire, the Impact of Event Scale-Revised (IES-R). PTSD diagnosis was clinically confirmed in all recovery (R-1, R-2) and maintenance (M-1, M-2) cohorts, with the Clinician-Administered PTSD Scale (CAPS-IV) administered in most participants.

### Reproducible latent dimensions across cohorts and both covariance matrix types

To characterize the dominant dimensions underlying trauma-related symptom variation, we first derived covariance matrices separately reflecting between-individual heterogeneity (hereafter referred to as between-individual covariance matrices) and within-individual temporal variation (hereafter referred to as temporal covariance matrices) within each longitudinal dataset (**Fig. 1A**). Principal component analysis (PCA) was then applied independently to each matrix. For between-individual covariance matrices, repeated observations were averaged within participant and symptom item to obtain a single symptom profile per individual. For temporal covariance matrices, participant-specific mean values were removed and residual symptom fluctuations were z-score normalized for each symptom item within each participant before concatenation across observations. Seven longitudinal cohorts each contributed one between-individual and one temporal covariance matrix, whereas two cross-sectional cohorts each contributed a between-individual covariance matrix only, yielding a total of 16 (= 7 x 2 + 2) covariance matrices.

**Fig. 1.**
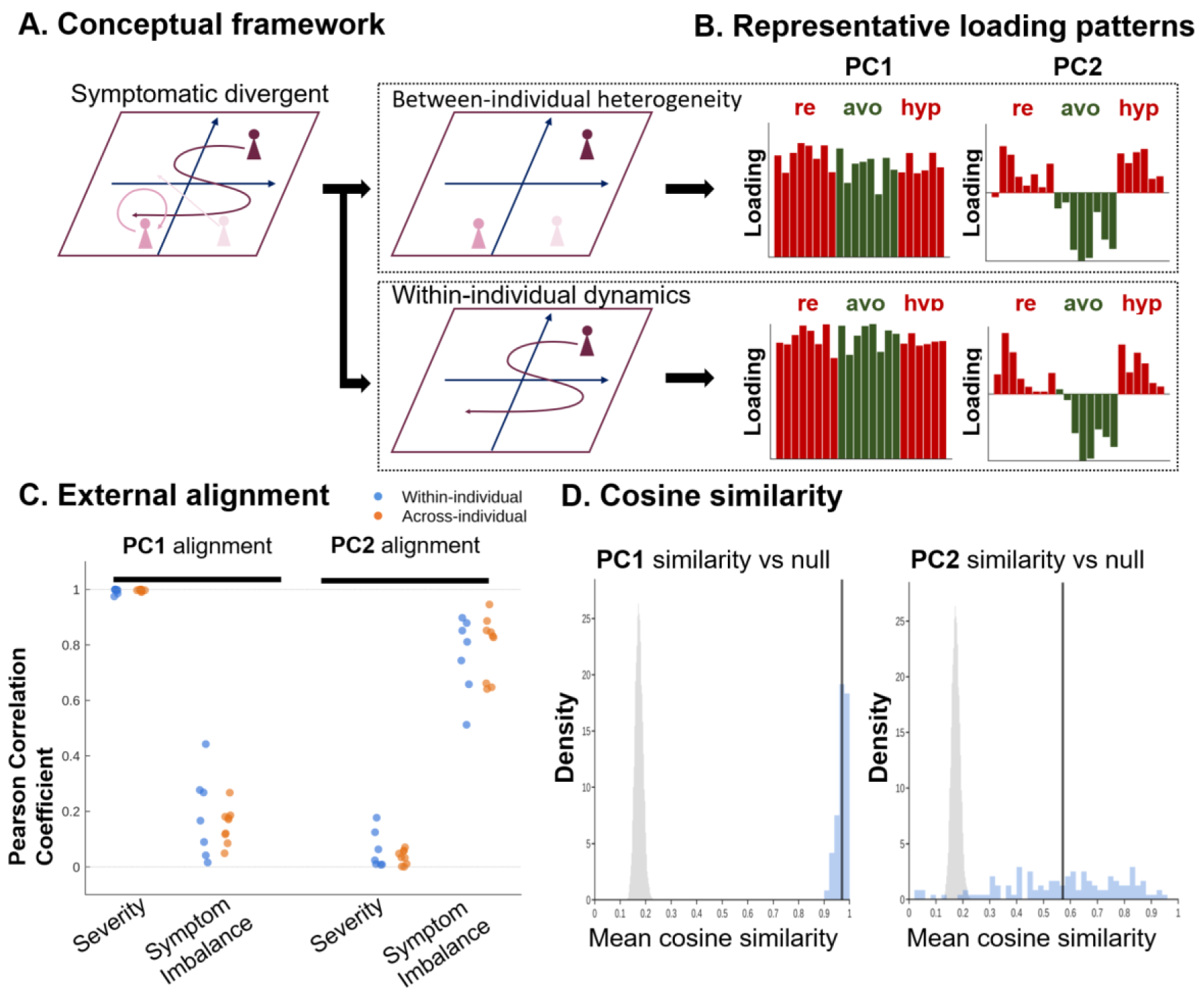
Reproducible shared latent dimensions across cohorts and both covariance matrix types. **A, Conceptual framework.** Trauma-related symptoms are represented in a low-dimensional state space capturing both between-individual heterogeneity and within-individual temporal variation. Individuals occupy distinct positions in this space (between-individual heterogeneity), while their symptom states evolve over time along trajectories within the same space (within-individual temporal variation). These symptom variabilities are hypothesized to be governed by shared latent dimensions that are consistent across cohorts and both covariance matrix types. **B, Representative loading patterns**. Representative loading patterns from one covariance matrix illustrating the first (PC1) and second (PC2) principal components. Representative loading patterns. RE denotes re-experiencing, AVO avoidance, and HYP hyperarousal. Red bars represent symptom clusters linked to emotional undermodulation, whereas green bars represent clusters linked to emotional overmodulation. Panel B shows a representative loading pattern from one covariance matrix for illustration. **C, External alignment.** Across covariance matrices, the first principal component (PC1) showed strong and selective alignment with overall symptom severity, whereas the second principal component (PC2) selectively aligned with the symptom imbalance index (re-experiencing score minus and avoidance scores). Each point corresponds to a temporal (blue) and between-individual (orange) covariance matrix. **D, Cosine similarity across covariance matrices.** The similarity of loading vectors across covariance matrices was evaluated using cosine similarity. The observed mean similarity for matched components (PC1–PC1 and PC2–PC2) is shown relative to a null distribution generated from randomly paired components across covariance matrices (PCx–PCy). Both PC1 (red line) and PC2 (blue line) exhibited substantially greater cross-covariance-matrix similarity than expected under the null model (gray histogram), indicating that the dominant symptom dimensions were consistently preserved across heterogeneous datasets.

Across all 16 covariance matrices, PCA consistently revealed two dominant latent dimensions (**Fig. 1B**). The first principal component (PC1) showed strong associations with overall symptom severity (Pearson’s r > 0.9 across datasets: **Fig. 1C** left). The second principal component (PC2) was strongly associated with the predefined symptom imbalance index (defined as the difference between re-experiencing and avoidance symptom scores) proposed in Chiba et al. (2020) (Pearson’s r > 0.5 across datasets: **Fig. 1C** right; also see **Quantitative variables**). Importantly, PC2 showed consistently weak associations with overall symptom severity across all covariance matrices (all |r| < 0.2), supporting its interpretation as a severity-independent axis. Across all pairwise comparisons among the 16 covariance matrices, loading patterns of the first two components were highly similar, with cosine similarity for matched components (PC1–PC1 and PC2–PC2) markedly exceeding a null distribution generated from randomly paired components (**Fig. 1D**). When analysis was restricted specifically to comparisons between between-individual and temporal covariance matrices, high cosine similarity was again observed, suggesting that shared latent dimensions underlie both between-individual heterogeneity and within-individual temporal covariation (**Supplementary Fig 1**).

We conducted several sensitivity analyses and confirmed that the main findings were robust to alternative temporal covariance estimation procedures (demeaning instead of z-score normalization; **Supplementary Fig. 2**), different symptom inclusion thresholds (primary analysis: IES-R ≥20; sensitivity analyses: IES-R ≥25 and ≥33; **Supplementary Fig. 3**), and alternative null distribution definitions for component similarity analyses (restricting null comparisons to the top 2 or top 5 principal components; **Supplementary Fig. 4**).

Because the first two principal components consistently captured overall symptom severity and the symptom imbalance dimension, subsequent analyses focused on these two components. To examine whether this focus could overlook additional reproducible dimensions, we next evaluated the stability of higher-order components across covariance matrices. Across all covariance matrix comparisons, loading similarity decreased progressively with increasing component order, with the first two principal components consistently showing the highest similarity across all comparison types (**Supplementary Fig. 5A**). In addition, the first two principal components together accounted for more than 50% of the variance in both between-individual and temporal covariance matrices (**Supplementary Fig. 5B**). Together, these findings suggest that the first two principal components capture the major reproducible dimensions across covariance matrices, justifying the primary focus on these components in subsequent analyses.

To examine whether PC2 reflected reproducible relationships among symptom clusters rather than an artifact of the PCA procedure, we analyzed severity-adjusted associations between symptom clusters. Across all cohorts and both covariance matrix types, severity-adjusted correlations between re-experiencing and avoidance symptoms were consistently negative and stronger in magnitude than those between re-experiencing and hyperarousal symptoms (**Fig. 2**). These findings provide an independent validation of the PC2 loading structure and indicate that the observed dimension reflects a reproducible pattern of symptom-cluster organization rather than a statistical consequence of PCA.

**Fig. 2.**
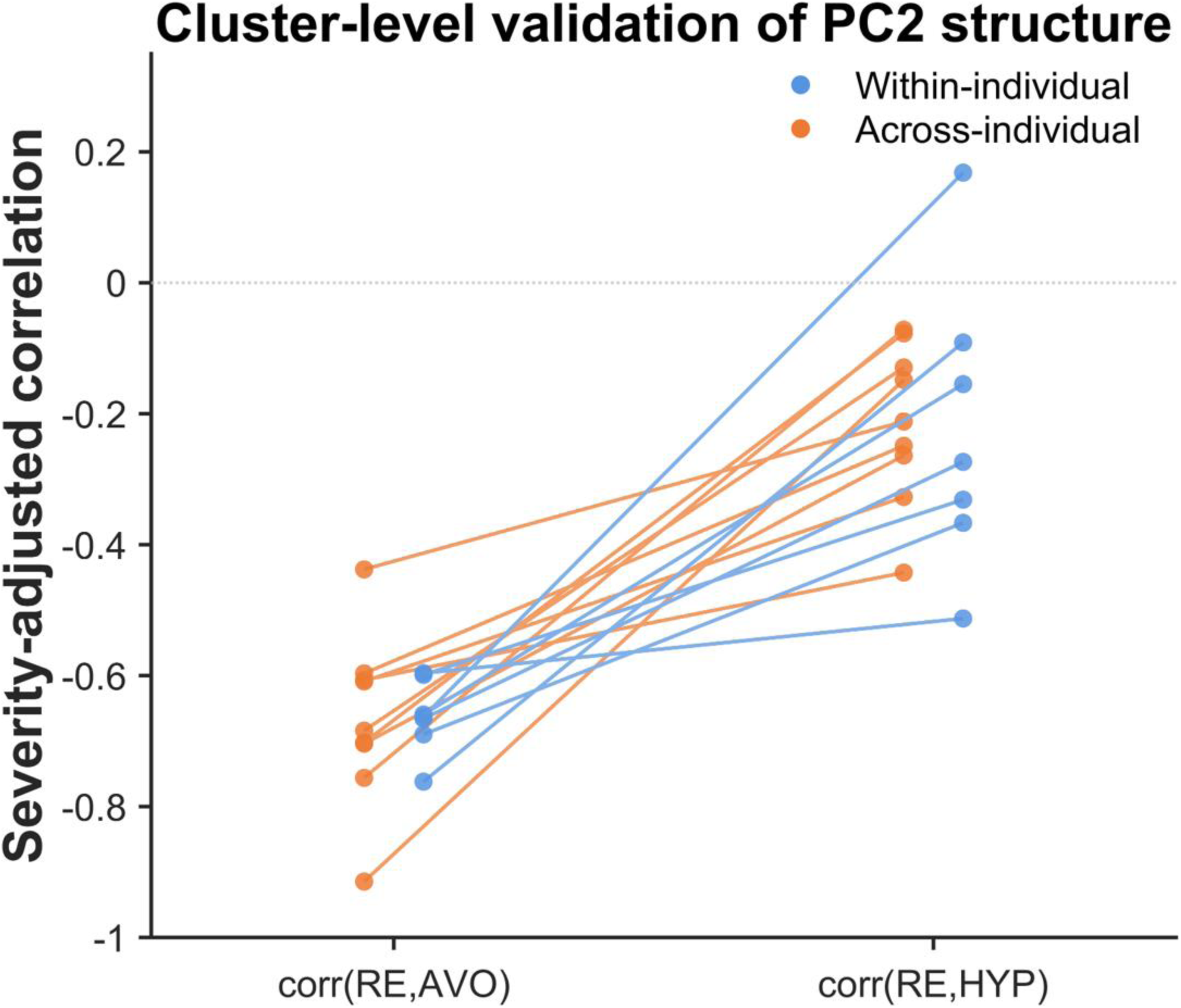
Severity-adjusted associations between PTSD symptom clusters. Severity-adjusted correlations between symptom clusters. Each line connects correlations derived from the same cohort and variance type. Re-experiencing (RE)–avoidance (AVO) severity-adjusted correlations were consistently more negative than re-experiencing (RE)–hyperarousal (HYP) correlations across cohorts and both between-individual and within-individual analyses. This pattern is consistent with our reciprocal inhibition model, in which RE and HYP are associated with emotional undermodulation, whereas AVO is associated with overmodulation. Importantly, these results do not imply that RE and AVO are negatively associated in absolute symptom severity, but rather that, after accounting for overall symptom severity, individuals differ in the relative dominance of RE versus AVO.

To assess whether the observed temporal covariance structure could arise trivially from concatenating repeated observations across individuals, we repeated PCA separately for each participant in two independent ultra-dense longitudinal cohorts (M-1–M-2). Components corresponding to symptom severity and symptom imbalance were reproduced in a substantial proportion of participants in both cohorts (**Supplementary Fig. 6**), supporting the presence of a similar temporal covariance structure at the individual participant level. Several permutation analyses demonstrated that the temporal covariance structure was not automatically derived from the between-individual covariance structure (**Supplementary Fig. 7**). Together, these findings indicate that the observed correspondence between between-individual heterogeneity and within-individual symptom dynamics is unlikely to reflect trivial artifacts arising from covariance estimation or data aggregation procedures.

### Universal symptom dimensions

Building on the observed cross-covariance-matrix similarity, we estimated shared symptom dimensions using hierarchical multi-group principal component analysis (hmgPCA). This approach yielded universal components capturing the shared latent dimensions across covariance matrices. As in the cohort-wise analyses, the first universal component (uPC1) corresponded to overall symptom severity, with broadly positive loadings across all symptom items. The second universal component (uPC2) captured the symptom imbalance dimension, characterized primarily by opposing contributions of re-experiencing and avoidance symptoms, with hyperarousal contributing in the same direction as re-experiencing (**Fig. 3A; Supplementary Table 2**). To evaluate the generalizability of these axes, we performed leave-one-out validation across covariance matrices. For each held-out covariance matrix, universal loading vectors estimated from the remaining covariance matrices were compared to covariance matrix-specific loading vectors using cosine similarity. Both uPC1 and uPC2 showed high cosine similarity across held-out covariance matrices, consistently exceeding null distributions generated from randomly paired components (**Fig. 3B-C**). Similar results were obtained when restricting the null distribution to the top two or top five principal components, indicating that generalizability was not driven by higher-order components **(Supplementary Fig. 8)**. These results demonstrate that the identified low-dimensional symptom axes are not only reproducible across covariance matrices and generalize to unseen covariance matrices.

**Fig. 3.**
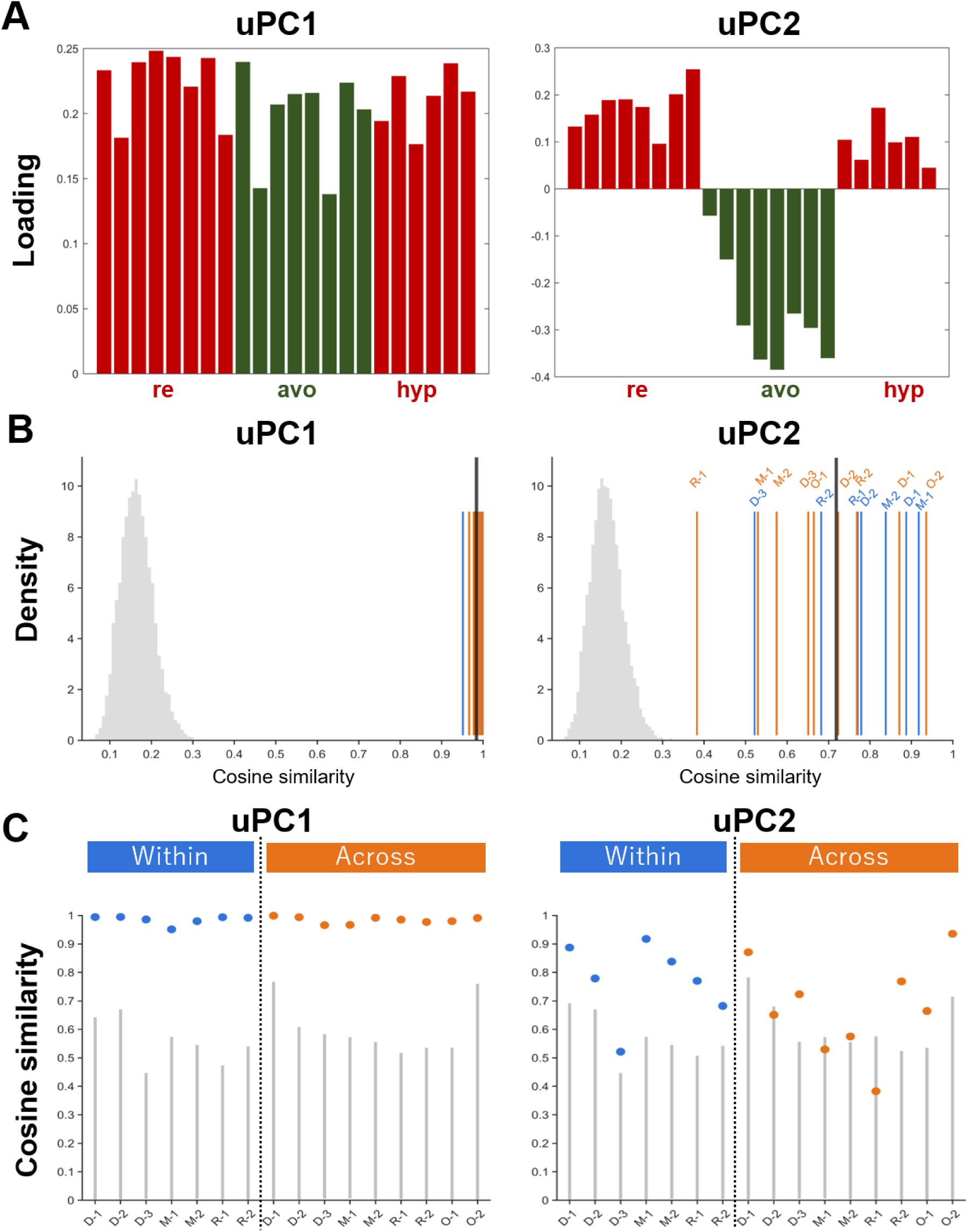
Universal symptom axes identified by hierarchical multi-group PCA. **A.** Loadings of the first two principal components (uPC1 and uPC2) obtained from hmg-PCA across all datasets. Each bar represents the loading of an individual symptom item. Symptoms are grouped by cluster, with re-experiencing (re), avoidance (avo), and hyperarousal (hyp) indicated above the corresponding items. **B.** Gray histograms show null distributions generated from randomly paired components. The black line indicates mean similarity across held-out covariance matrices for PC1 (left) and PC2 (right), while blue and orange lines represent similarity values for individual within-individual and between-individual covariance matrices, respectively. **C.** Cosine similarity for each held-out covariance matrix is shown alongside the corresponding null distribution (95% interval). Each point represents the observed similarity for a single covariance matrix, and vertical lines indicate the range of values obtained under random component pairings.

### Temporal variation

We quantified the relative magnitude of within-individual temporal variation along uPC2 by computing, for each participant, the ratio of within-individual variance to cohort-level between-individual variance. In 32.9% of participants (5,110 of 15,549), within-individual variability exceeded between-individual variability (variance ratio > 1), with similarly long-tailed distributions observed across cohorts (**Fig. 4; Supplementary Fig. 9**). The median variance ratio was 0.58 (IQR 0.21–1.34), indicating that variability along uPC2 is not dominated by stable between-individual heterogeneity but also reflects pronounced within-individual temporal variation. Analyses were restricted to participants who exceeded the clinical cutoff (IES-R ≥ 20) at least once. When all participants were included, the proportion of individuals with variance ratios > 1 was lower (∼25%), but the overall distributional pattern remained qualitatively unchanged (**Supplementary Fig. 10**).

**Fig 4.**
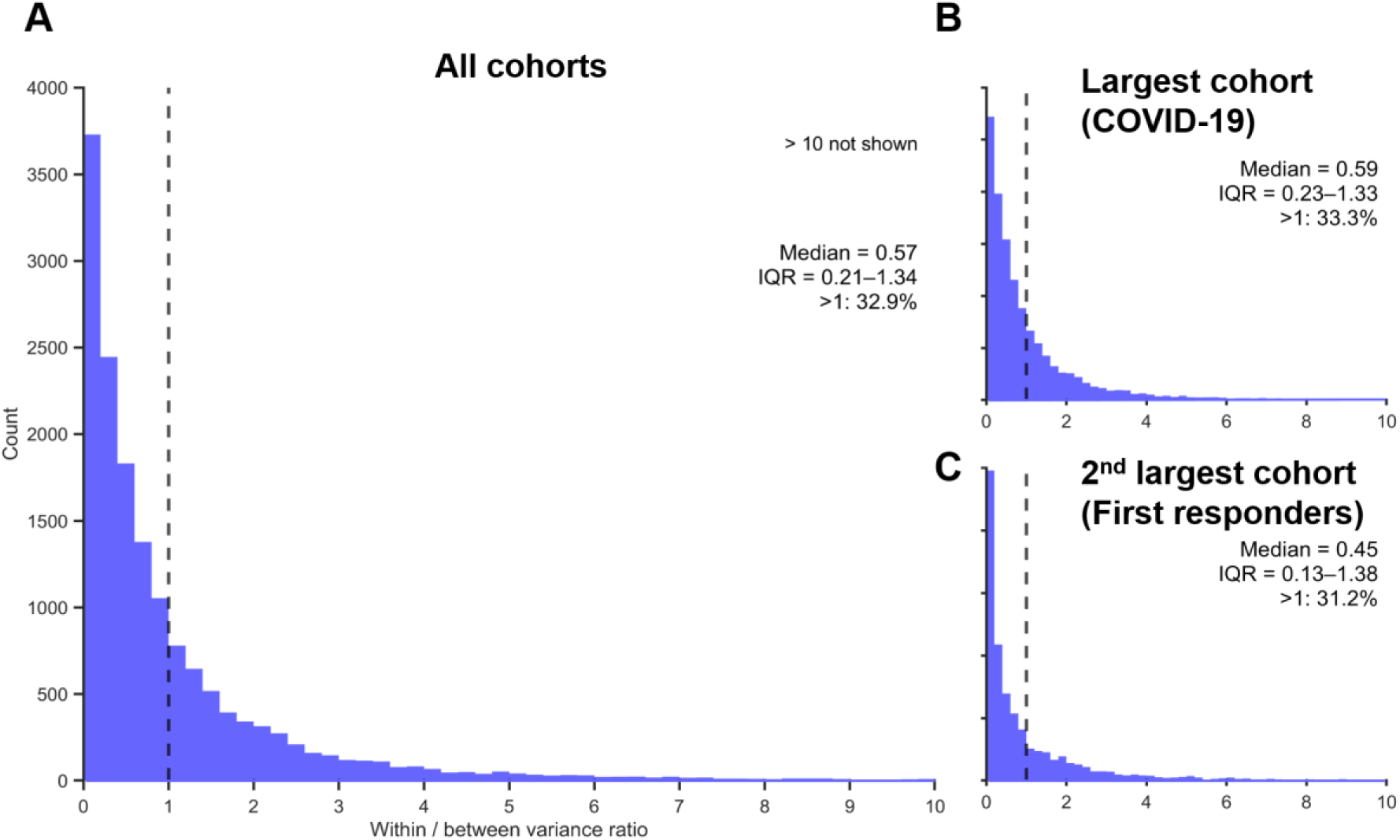
The distribution of within-to-between variance ratios along uPC2. **(A)** Distribution across all cohorts. **(B–C)** Distributions for the largest (COVID-19) and second-largest (First responders) cohorts. Distributions are centered below or around unity but exhibit a pronounced right tail, indicating that while variability is partly constrained by stable between-individual heterogeneity, a subset of individuals shows strong within-individual temporal variation. The dashed line indicates a ratio of 1. Values >10 are not shown for visualization but are included in statistical analyses.

### Treatment trajectories

We compared symptom trajectories from CPT and EMDR cohorts (i.e. R-1 and R-2) along the universal axes using within-individual pre–post changes (**Fig. 5A**). To maximize available treatment data, missing item values were conservatively imputed as described in Methods, which recovered data from three additional CPT participants. After excluding participants without recoverable pre–post observations, treatment analyses included 16 participants in the CPT cohort and 20 participants in the EMDR cohort. Both treatments produced comparable reductions along the severity axis (uPC1), with no significant difference between groups (*t* = -1.31, *df* = 34, *p* = 0.20; **Fig. 5B**). In contrast, CPT and EMDR cohorts showed mean changes in opposite directions along uPC2 (*t* = -3.04, *df* = 34, *p* = 0.005; **Fig. 5C**), with CPT increasing uPC2 and EMDR decreasing it. These opposite shifts suggested relatively greater reductions in avoidance symptoms following CPT and in re-experiencing symptoms following EMDR. No significant difference in uPC2 was observed at baseline (mean difference = 1.16, *t* = 1.89, *df* = 34, *p* = 0.067). The relative positions of the CPT and EMDR cohorts along uPC2 reversed over the course of treatment, indicating that treatment-related divergence along uPC2 emerged during treatment rather than reflecting pre-existing group differences. Together, these findings indicate that the observed divergence in uPC2 was not explained by differences in baseline severity or baseline position along the same axis. No between-group differences were observed along uPC1 at either baseline (*t* = 0.42, *df* = 34, *p* = 0.68) or post-treatment (*t* = -0.93, *df* = 34, *p* = 0.36). Moreover, the treatment effect on ΔuPC2 remained significant after controlling for baseline uPC1 and uPC2 using ANCOVA (*β* = -1.11, *p* =0.03). Related treatment differences were also observed using supplementary analyses based on conventional symptom-cluster measures (**Supplementary Fig. 11**). Sensitivity analyses restricted to complete observations yielded qualitatively similar results overall, although the ANCOVA effect on ΔuPC2 was attenuated in the complete-case analysis and fell slightly below the conventional significance threshold (*p* = 0.087; **Supplementary Table 3**). Overall, both treatment cohorts were predominantly composed of participants with interpersonal trauma, and no major medication changes were identified during treatment (see **Table 1**).

**Fig 5.**
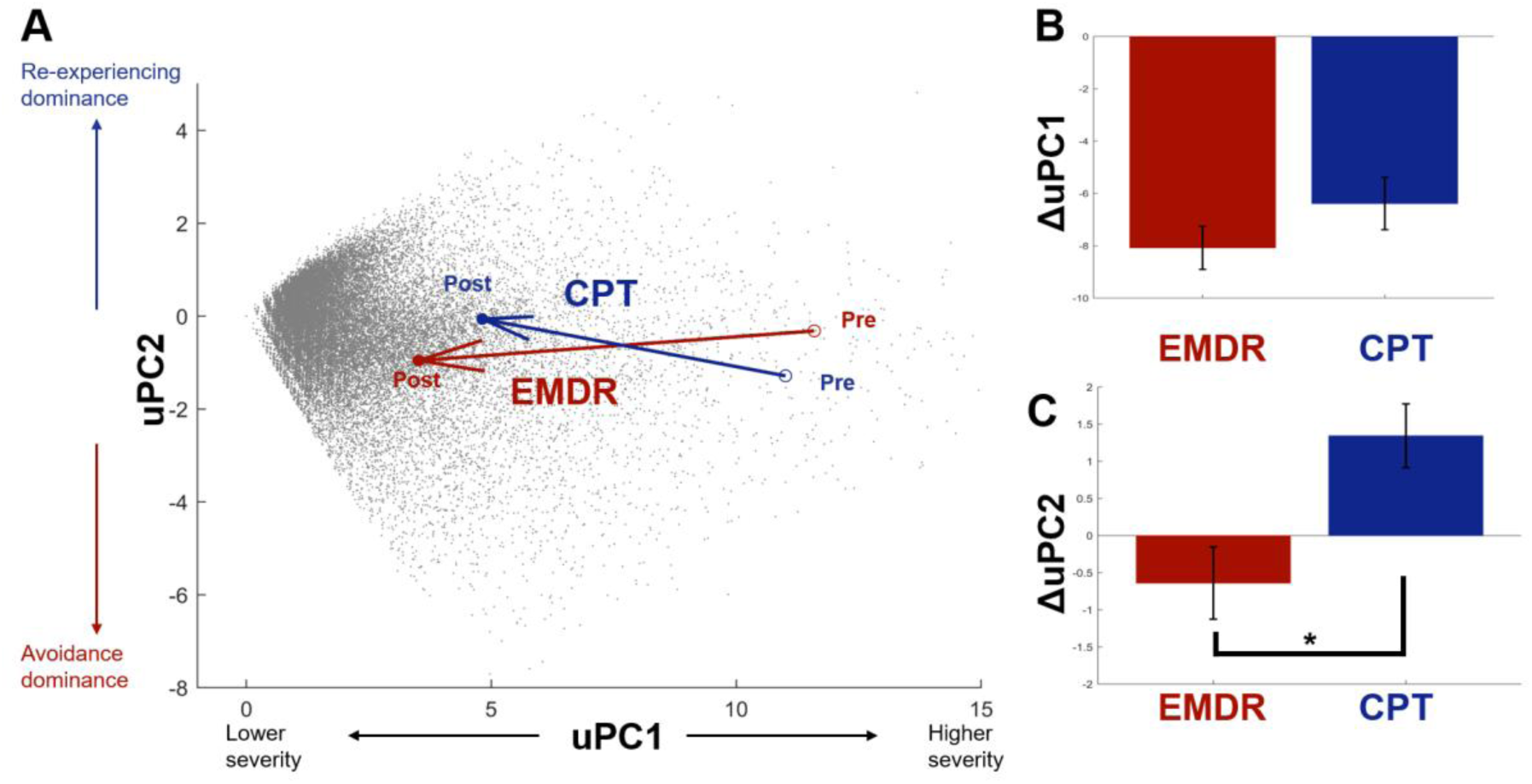
CPT and EMDR shift patients in opposite directions along the symptom imbalance dimension despite comparable reductions in symptom severity. **A.** Group-level trajectories of cognitive processing therapy (CPT: R-1 in **Table 1**; blue) and eye movement desensitization and reprocessing (EMDR: R-2 in **Table 1**; red) in the universal symptom space defined by uPC1 (severity) and uPC2 (symptom imbalance). Arrows represent group-level mean changes from pre-treatment (open-circles) to post-treatment (filled circles). Higher and lower uPC2 values indicate relatively greater re-experiencing and avoidance symptom dominance, respectively. B. Pre–post changes along uPC1. Both treatments reduced symptom severity to a similar extent, with no significant difference between groups. C. Pre– post changes along uPC2. In contrast to uPC1, CPT and EMDR showed opposite mean changes along uPC2, indicating treatment-related divergence that was not apparent from overall symptom severity alone. Gray points in the background indicate individual observations from the Japanese Self-Defense Force cohort (D-1 in **Table 1**), shown to illustrate the overall distribution of symptom states within the universal symptom space. Error bars represent standard error of the mean. \**p* < 0.05.

## DISCUSSION

This study demonstrates that trauma-related symptom heterogeneity, spanning both between-individual and within-individual temporal variation, is captured by shared latent dimensions. These dimensions comprised a first component reflecting overall symptom severity and a second severity-independent component capturing variation primarily in the relative contribution of re-experiencing and avoidance symptoms. The shared latent dimensions were reproducible across multiple cohorts spanning distinct clinical phases, including development, chronicity, and recovery, regardless of whether symptom variation reflected between-individual heterogeneity or within-individual temporal variation. These shared dimensions were summarized as universal axes (uPC1 and uPC2), which enabled direct comparison of symptom trajectories across cohorts and treatments. Notably, we found that two treatment approaches commonly used for PTSD (CPT and EMDR) exhibited distinct effects in opposite directions along uPC2, with relatively greater reductions in avoidance or re-experiencing symptoms, respectively.

The uPC2 captured symptom imbalance, a dimension previously introduced to operationalize theories of emotional under- and overmodulation in PTSD. These theories describe PTSD symptom expression as varying along a continuum between insufficient and excessive emotional modulation. The emergence of a shared symptom imbalance dimension across both between-individual heterogeneity and within-individual temporal variation raises the possibility that both forms of symptom variation may reflect a common emotion-regulation process. This finding provides empirical support for a key conceptual assumption underlying the reciprocal inhibition model, which posits that both between-individual heterogeneity and within-individual dynamics arise from the same underlying regulatory process involving dynamic shifts in relative dominance between the amygdala and vmPFC^26^. However, the present analyses do not directly test the psychological or neurobiological mechanism underlying this dimension. Future work can investigate the neurobiological processes underlying variation along this dimension, including amygdala-vmPFC circuit dynamics.

The uPC2 may have clinical relevance for at least two reasons. First, variability along uPC2 frequently reflected substantial within-individual temporal variation, indicating that symptom variation along this dimension may represent a clinically meaningful process within individuals. Second, uPC2 may capture clinically meaningful differences between two first-line psychotherapies for PTSD—CPT and EMDR, calling for a state dependent treatment. Both interventions produced comparable improvements along uPC1, but diverged systematically along uPC2. Specifically, CPT shifted symptom trajectories toward higher uPC2 values, corresponding to relatively greater reduction in avoidance symptoms than re-experiencing symptoms, whereas EMDR shifted trajectories in the opposite direction. This CPT-related shift aligns with therapeutic frameworks emphasizing cognitive processing of trauma-related memories and reduced reliance on avoidance-related processing. In contrast, the EMDR-related shift may reflect the distinctive emphasis of EMDR on lower-level sensory-affective processing through bilateral stimulation, a core component of EMDR, rather than primarily on cognitive restructuring. This interpretation is consistent with experimental evidence that attenuates amygdala activity^27^, a region strongly implicated in re-experiencing symptoms^24,28^. These findings are broadly consistent with previous meta-analytic evidence that EMDR may produce relatively greater improvement in re-experiencing symptoms relative to other trauma-focused psychotherapies^29^, including CPT, despite comparable effects on overall severity^30^. Together with substantial within-individual temporal variation along uPC2, these findings raise the possibility that differential treatment response may depend partly on an individual’s current position along this symptom dimension. In particular, EMDR and CPT may engage partially distinct symptom processes related to relatively greater re-experiencing and avoidance states, respectively. Notably, conventional symptom-cluster analyses alone did not reveal that these treatment differences were organized along a severity-independent symptom dimension (**Supplementary Fig. 11**), highlighting the added value of the present low-dimensional framework. Although prospective validation is needed, such dimensions may help generate rational hypotheses for treatment selection in settings where definitive comparative evidence is lacking.

The same latent dimensions were observed not only in clinically diagnosed PTSD cohorts but also in non-clinical cohorts capturing stress responses to the COVID-19 pandemic, and remained stable across lower IES-R thresholds. This broader generalizability suggests that the identified dimension may reflect a more general organizing principle of stress-related cognitive variability rather than a process specific to PTSD or trauma exposure. This possibility may extend beyond trauma-related symptoms, as partial overlap between within-individual dynamics and cross-sectional symptom organization has also been reported in disorders including depression and schizophrenia^19,21^. Thus, current dimensional psychiatry frameworks such as RDoC and HiTOP may need to be expanded to explicitly incorporate within-individual dynamics over time. This may enable knowledge accumulated from between-individual heterogeneity—including symptom dimensions, neurobiological correlates, and treatment-related markers—to inform the study of symptom dynamics over time. For example, previous studies have linked symptom dimensions and neurobiological subtypes to differential treatment response, including functional-connectivity-based depression biotypes associated with TMS outcomes^6^ and prefrontal-amygdala circuitry associated with psychotherapy response in PTSD^9^. Since prefrontal-amygdala circuitry overlaps with the neural circuit hypothesized to underlie uPC2, an important future direction will be to examine whether temporal variation in this circuit maps onto symptom dynamics along uPC2 and thereby contributes to differential psychotherapy response. More broadly, psychiatric heterogeneity may not be fully understood as a collection of static subtypes, but may also be represented as trajectories within a common low-dimensional symptom space. This perspective raises the possibility that systematic within-individual dynamics may contribute to the challenges of reproducing some symptom- and circuit-based subtypes across cohorts^12,13^. More fundamentally, these findings suggest that psychiatric intervention may need to be conceptualized not only in terms of matching treatments to patient subtypes, but also in terms of dynamically matching interventions to an individual’s current symptom state. This perspective highlights the importance of delivering the right treatment at the right time, and suggests that interventions designed to shift patients toward more treatment-responsive states could enhance the efficacy of subsequent therapies.

One possibility is that the second component reflects statistical properties of the measurement instrument rather than a meaningful dimension. However, the high reproducibility of similar loading structures across heterogeneous cohorts, together with the failure of permutation analyses to reproduce comparable covariance structures, argues against a purely instrument-driven or dataset-specific artifact and instead supports the presence of a robust covariance structure across symptom clusters. Consistent with this interpretation, severity-adjusted correlations between re-experiencing and avoidance symptoms were consistently stronger than those between re-experiencing and hyperarousal symptoms across cohorts and variance types, independently supporting the validity of the uPC2 loading structure. Together, these findings suggest that symptom imbalance dimension possesses several properties expected of a low-dimensional state variable, including reproducibility across populations, organization of both within- and between-individual variation, and systematic modulation by therapeutic interventions. This raises the hypothesis that symptom imbalance may function as a candidate order parameter for PTSD dynamics, providing a basis for future studies testing whether spontaneous fluctuations along this dimension are quantitatively related to responses to therapeutic perturbations^31–37^.

Several limitations should be considered. First, symptom measurement was based on a single self-report instrument (IES-R). While the IES-R largely overlaps with DSM-based and other trauma-related measurements, important differences remain in symptom coverage, suggesting that comparison or integration with broader assessments may reveal additional symptom dimensions beyond those identified here. Second, PCA captures linear covariance structures and may not fully represent nonlinear relationships between symptoms; nonlinear manifold learning approaches may provide complementary insights into the geometry of symptom variation^38^. Third, the present findings identify reproducible symptom dimensions but do not establish the underlying mechanisms that generate variation along these dimensions. Although the present framework was motivated by amygdala-vmPFC reciprocal inhibition, future studies integrating broader large-scale network models, such as triple-network models^39,40^, may help determine whether similar low-dimensional symptom organization reflects more general principles shared across psychiatric disorders. Finally, CPT and EMDR were derived from independent cohorts rather than being directly compared within a randomized controlled design. Therefore, the observed treatment-related differences may partly reflect cohort-specific characteristics, selection bias, therapist-related factors, or other unmeasured confounding variables rather than treatment-specific mechanisms. Prospective controlled studies will be required to establish causal effects of different psychotherapies on symptom trajectories, which may subsequently inform whether treatment can be optimized through state-dependent intervention.

In summary, trauma-related symptom heterogeneity—between individuals and within individuals over time—is captured by shared latent dimensions corresponding to overall severity (uPC1) and severity-independent symptom imbalance (uPC2). Within this framework, CPT and EMDR showed comparable improvements along the severity dimension (uPC1) but divergent trajectories along the symptom imbalance dimension (uPC2), revealing treatment differences that are not visible when outcomes are summarized solely by total symptom scores. More broadly, these findings suggest that psychiatric heterogeneity may need to be understood not only in terms of where patients are located within symptom space, but also how they move through that space over time. This perspective opens the possibility of a more temporally informed precision psychiatry in which interventions are tailored not only to the individual, but also to the patient’s current symptom state and trajectory of recovery.

## METHODS

### Study design

To test whether symptom imbalance emerges as a shared dimension across both between-individual heterogeneity and within-individual temporal variation, we conducted principal component analysis (PCA) separately on between-individual variation (mean differences) and within-individual variation (demeaned and standardized temporal variations) using seven longitudinal datasets and two large cross-sectional online datasets, thereby enabling independent evaluation of their covariance structures. After identifying latent dimensions separately for each cohort and each source of variation, we integrated the corresponding covariance matrices using hierarchical multi-group PCA to estimate universal axes (uPC1/uPC2). Finally, to examine the clinical relevance of the identified universal axes, symptom trajectories from psychotherapy cohorts were projected onto this shared symptom space to quantify treatment-related trajectories within it.

### Setting

This study integrated seven longitudinal and two cross-sectional cohorts capturing trauma-related symptoms across different clinical phases, including development, maintenance, and treatment-related recovery. Cohort characteristics, recruitment procedures, and assessment schedules are summarized in **Table 1** and **Supplementary Methods**.

All studies were conducted in accordance with the Declaration of Helsinki and were approved by the institutional review boards of the participating institutions, including the National Defense Medical College, Ground Defense Force Hanshin Hospital, National Center of Neurology and Psychiatry, and Advanced Telecommunications Research International.

### Participants

Participants were adults who had experienced traumatic or highly stressful events and completed the IES-R. Because the study aimed to examine shared symptom dimensions across a broad range of trauma- and stress-related responses, cohorts were not restricted to individuals meeting DSM-defined PTSD Criterion A or formal PTSD diagnoses. Participants were included if they exceeded the symptom cutoff (IES-R ≥20) at least once. Between-individual analyses included all available observations from eligible participants, whereas within-individual analyses were restricted to participants with at least three repeated assessments required for covariance estimation (**Supplementary Methods**).

### Variables

PTSD-related symptoms were assessed using the IES-R^41,42^, a 22-item self-report questionnaire measuring trauma-related distress across re-experiencing, avoidance, and hyperarousal clusters. Each item is rated on a five-point Likert scale (0–4), yielding a total score range of 0–88. The Japanese version of the IES-R has been validated for reliability and construct validity^41^ and was consistently used across the cohorts analyzed in this study.

To reduce floor effects while retaining clinically meaningful symptom variation, an IES-R cutoff of ≥20 was used in the main analyses^41,43^. Sensitivity analyses were conducted using higher thresholds (≥25 and ≥33)^41,42^.

Compared with other DSM-based measures such as the PTSD Checklist (PCL) and the Clinician-Administered PTSD Scale (CAPS), the IES-R largely overlaps in assessing re-experiencing, avoidance, and hyperarousal symptoms, but does not include most symptoms from the DSM-5 cluster of negative alterations in cognition and mood (NACM). Notably, both re-experiencing and most avoidance items in the IES-R explicitly reference the traumatic event itself (“IT” in **Supplementary Table 2**), including symptoms such as emotional numbing and dissociative responses that are currently categorized separately within DSM-5. In contrast, hyperarousal and NACM symptoms are generally not explicitly anchored to the traumatic event. Overall, the IES-R places greater emphasis on trauma memory-related symptomatology, in line with the original rationale for defining symptom imbalance along the re-experiencing–avoidance dimension.

### Data measurement

Assessment intervals varied substantially across cohorts, ranging from every 5 days to annual follow-up assessments (**Table 1**). Because temporal sampling frequency differed substantially across cohorts, analyses focused on covariance structures of symptom variation rather than temporal dynamics defined by absolute sampling intervals.

### Study size

Sample size reflected the total availability of eligible existing datasets rather than prospective power calculations. The full dataset (before applying analytic inclusion criteria) included 100,077 participants and 327,800 observations across the nine cohorts. Cohort sizes ranged from 12 to 51,551 participants (median = 68). Because the primary objective was to estimate stable covariance structures across heterogeneous cohorts rather than test a predefined effect size, all eligible observations were retained for analysis.

### Quantitative variables

Longitudinal symptom data were decomposed into complementary components of variation for each symptom item within each individual. Between-individual variation was estimated as each participant’s mean symptom profile across time points. Within-individual variation was estimated by z-scoring each symptom time series within individuals, thereby removing between-person mean differences and normalizing variance across participants. This approach enabled covariance structures to reflect relative symptom co-fluctuation rather than absolute severity differences, while minimizing potential mean–variance coupling whereby individuals with higher average symptom severity may exhibit inflated temporal variance. As a sensitivity analysis, within-individual variation was alternatively estimated using demeaned time series.

To facilitate interpretation of principal components, the symptom imbalance index was defined a priori as the difference between re-experiencing and avoidance symptom clusters following our previously proposed emotion modulation framework^26^. This definition specifically focuses on trauma-specific symptom expression, as re-experiencing and avoidance symptoms are directly defined by their relationship to trauma-related memories and cues, whereas hyperarousal symptoms primarily reflect more general stress-related responses. Higher values indicate relative dominance of re-experiencing symptoms, whereas lower values indicate relative dominance of avoidance symptoms. This index was used solely as an interpretive framework and was not used in PCA estimation.

### Statistical methods

We used principal component analysis (PCA) as a descriptive dimension-reduction approach to summarize dominant covariance patterns among observed symptoms. Our aim was not to infer latent psychological constructs underlying symptoms, as typically emphasized in factor-analytic models, but rather to identify reproducible axes of symptom variation across cohorts. Accordingly, the identified components should be interpreted primarily as empirical axes of symptom variation, while their underlying psychological interpretation remains to be established. All analyses were conducted using MATLAB 2021a (MathWorks). Statistical significance was assessed using two-sided tests unless otherwise specified.

### Cohort-specific principal component structure

Within each cohort, PCA was performed separately on symptom-level covariance matrices derived from between-individual heterogeneity and within-individual variation. The first two principal components (PC) were retained because they consistently captured the dominant covariance patterns across cohorts and showed reproducible loading structures with consistent clinical interpretability. To facilitate component interpretation, Pearson correlations were calculated between PC1/PC2 scores and 1) overall symptom severity (IES-R total score) and 2) the predefined symptom imbalance index (see **Quantitative variables**). Structural similarity across cohorts and variance types was quantified using cosine similarity between L2-normalized loading vectors, using absolute similarity values to account for the arbitrary sign indeterminacy. Statistical significance was assessed against a null distribution generated by randomly pairing PCs across cohorts (PCx–PCy). To ensure comparability across cohorts, the primary analysis used the minimum number of estimable principal components across datasets (11 components). Sensitivity analyses evaluated alternative null distributions and component pairing schemes. Additional analyses tested whether the severity (SEV) and symptom imbalance (SI) dimensions could be recovered from PCA performed separately for individual participants **(Supplementary Methods).**

### Severity-adjusted cluster associations

To assess whether the secondary symptom dimension reflected reproducible relationships among symptom clusters rather than a consequence of PCA decomposition alone, we examined severity-adjusted associations between IES-R symptom clusters. Mean re-experiencing, avoidance, and hyperarousal scores were computed using standard IES-R subscale definitions. For between-individual analyses, participant-level mean subscale scores were calculated across all available observations. For within-individual analyses, subscale scores were analyzed separately within each participant. Within each participant, re-experiencing, avoidance, and hyperarousal scores were individually regressed on total IES-R score, and residual scores were retained. Residual scores were then pooled across participants for correlation analyses. Pearson correlations were then calculated between residualized re-experiencing and avoidance scores and between residualized re-experiencing and hyperarousal scores. Correlations were computed separately for between-individual and within-individual covariance structures.

### Hierarchical multi-group principal component analysis

To identify shared symptom dimensions across cohorts, hierarchical multi-group principal component analysis was applied to cohort-specific covariance matrices. For each cohort, covariance matrices were estimated separately for between- and within-individual variation (see **Quantitative variables**). Rather than pooling raw observations across cohorts, covariance matrices were first estimated within each cohort and subsequently integrated. This approach prevented large cohorts and specific clinical phases from disproportionately shaping the resulting shared dimensions, thereby preserving balanced contributions across cohorts and variance types. Cohort-level covariance matrices were aggregated by averaging across cohorts without weighting by sample size so that each cohort contributed equally to the estimation of the shared dimensions. Eigenvalue decomposition of the aggregated covariance matrix was performed to estimate universal principal components (uPC1 and uPC2), representing the shared principal axes across covariance matrices.

Robustness of the universal axes was evaluated using leave-one-out validation across covariance matrices. For each loading vector from the held-out covariance matrix, cosine similarity was computed with universal loading vectors estimated from the remaining cohorts. Mean similarity across held-out covariance matrices was used as the primary summary metric. Similarity values for individual covariance matrices were additionally examined to assess variability in generalization performance. Statistical significance was assessed using permutation-based null distributions generated from randomly paired components (PCx–PCy). Alternative null-model specifications were evaluated in sensitivity analyses (Supplementary Methods).

### Relative magnitude of within-individual temporal variation along uPC2

To quantify the relative magnitude of within-individual temporal variation compared with between-individual heterogeneity, variance along the universal second principal component (uPC2) was calculated separately for within-individual variation for each individual and for cohort-level between-individual variation. A variance ratio was computed for each individual as the ratio of within-individual variance to cohort-level between-individual variance. The distribution of variance ratios was summarized using histograms and descriptive statistics, and individuals for whom the ratio exceeded one were counted.

### Projection of treatment trajectories

To evaluate clinical relevance, symptom trajectories from psychotherapy cohorts were projected onto the universal dimensions. For treatment-effect analyses, missing item values were conservatively imputed when no more than one item was missing within a given symptom cluster, by replacing the missing value with the mean of the remaining items from the same symptom cluster, participant, and time point. Symptom vectors were multiplied by the universal loading matrix to obtain uPC scores for each observation. Treatment effects were quantified as within-individual changes in uPC scores between pre- and post-treatment observations. Differences in change between treatment cohorts were tested using two-sample t-tests. To examine the effects of baseline imbalance, baseline uPC scores were compared between treatment cohorts. In addition, treatment-related changes in uPC2 were evaluated using ANCOVA, with treatment group as the predictor and baseline uPC1 and baseline uPC2 scores included as covariates. As a sensitivity analysis, all treatment-effect analyses were repeated using complete observations only, without imputation.

## Data availability

The datasets analyzed in the current study contain sensitive clinical and personal information and cannot be made publicly available because of restrictions imposed by the ethics committees and institutional review boards that approved the studies. De-identified data can be made available from the corresponding author upon reasonable request, subject to institutional approval and applicable data-sharing agreements. The aggregated covariance matrix used for hierarchical multi-group PCA, together with the universal component loadings (uPC1 and uPC2) required to project new observations into the symptom space, will be made publicly available upon publication.

## Code availability

Custom MATLAB code used for data preprocessing, principal component analyses, hierarchical multi-group PCA, and statistical analyses will be made publicly available upon publication.

## Acknowledgements

This study is supported by JSPS KAKENHI (JP24K18731), AMED (JP24wm0625502), the Japan Society for the Promotion of Science (JSPS) (23K24266; HT), the National Defense Medical College (NDMC) (55; HT), the KDDI collaborative research contract, and the Institute for Basic Science (IBS-R015-D2; AC). Petra Ritter acknowledges support from the EU4Health Programme through SCAILED (101314097); the EU Horizon Europe Programme through BRIDGE (101219311), EBRAINS2.0 (101147319), Virtual Brain Twin for Personalized Treatment of Psychiatric Disorders (101137289), EBRAINS-PREP (101079717), AISN (101057655), EBRAIN-Health (101058516), and EIC grant PHRASE (101058240); the Digital Europe Programme through TEF-Health (101100700), SHAIPED (101195135), and CoordinaTEF (101168074); the German Research Foundation (DFG) through SFB 1436 (Project ID 425899996), SFB 1315 (Project ID 327654276), SFB 936 (Project ID 178316478), SPP Computational Connectomics (RI 2073/6-1, RI 2073/10-2, and RI 2073/9-1), DFG Clinical Research Group BECAUSE-Y (504745852), the DFG German-Spanish project BrainsAge (RI 2073/14-1), and DFG Project ID 424778381 – TRR 295 Retune; ERA PerMed JTC2021; the German Federal Ministry for Health through PatternCog (ZMI5-2522FSB904); Berlin University Alliance OpenMake; and the Berlin Institute of Health and Foundation Charité. The funders had no role in the study design, data collection and analysis, the decision to publish, or in preparation of the manuscript. We thank Miho Nagata for administrative and research coordination support.

## Author Contributions

T.C. conceived and designed the study, performed the main analyses, interpreted the results, and wrote the manuscript. M.It. and M.Ic. contributed to data collection, interpretation of the findings, scientific discussion, and manuscript preparation. M.M. contributed to data collection across multiple cohorts, performed data analyses, and contributed to interpretation of the results. M.N., T.Sa., T.Te., H.T., N.K., M.S., K.I., H.K., M.H., K.N., and T.Ka. contributed to data collection, cohort management, and critical review of the manuscript. A.C., E.V., E.J.G., F.H.S.v.d.D., N.J.v.d.W., N.G., M.K., P.R., T.Ki., T.Ku., and T.H. contributed to scientific discussion, interpretation of findings, conceptual refinement, and critical revision of the manuscript. All authors read and approved the final manuscript.

