## Supplementary Material for "Empirical validation of a predicted emotional modulation dimension linking temporal variability and individual differences in PTSD"

**Supplementary Methods**

Basic demographic characteristics and sample sizes for all cohorts prior to analytic exclusion are summarized in **Supplementary Table 1**. Detailed descriptions of individual cohorts are provided below.

**Supplementary Table 1 Full cohort characteristics and sample sizes**

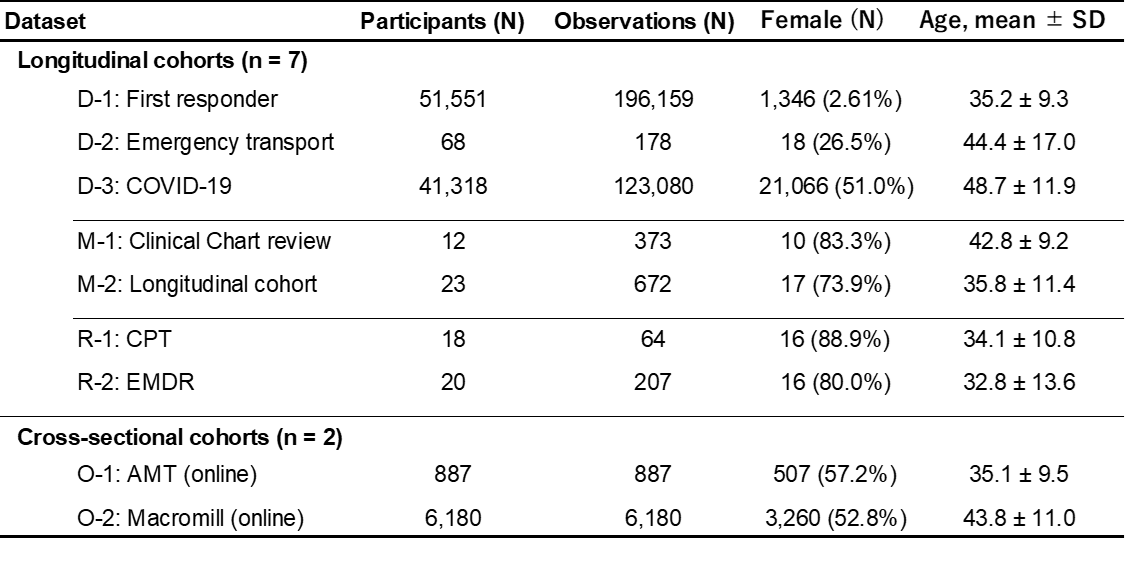

**D-1** **(First responder cohort):** Participants were Japan Self-Defense Forces personnel deployed to disaster-relief operations following the 2011 Great East Japan Earthquake. Assessments were administered annually between 2013 and 2018. Cohort details have been described previously^1,2^.

**D-2 (Emergency transport cohort)**: Participants were patients transported to the National Defense Medical College Hospital following traffic-related trauma meeting DSM PTSD Criterion A. The present study used longitudinal symptom data collected as part of a larger prospective trauma study. Data collection occurred between 2021 and 2024. Because the initial assessment was conducted within 48 hours after trauma exposure, participants were instructed to report their current symptom state rather than symptoms experienced over the preceding week specified in the original IES-R instructions.

**D-3** **(COVID-19 online cohort):** Participants were recruited from the general population through the Macromill online survey platform during the COVID-19 pandemic. Surveys were conducted across five waves between August 2020 and December 2021. Cohort details have been described previously^3^.

**M-1** **(Clinical chart-review cohort):** Participants were outpatients with PTSD who completed the IES-R during routine clinical visits. Data collection occurred between March 2014 and September 2018.

**M-2** **(Longitudinal cohort):** Participants met diagnostic criteria for PTSD and were recruited from Ground Defense Force Hanshin Hospital and Seven Mental Clinic. Assessments were administered every 5 days for 6 months using smartphone-based questionnaires. Data collection occurred between April 2025 and December 2025. This cohort was established specifically for the present study to obtain high-frequency longitudinal symptom measurements. Because assessments were conducted at 5-day intervals, participants were instructed to report symptoms experienced since the previous assessment rather than during the preceding week specified in the original IES-R instructions.

**R-1** **(Cognitive Processing Therapy cohort)**: Participants met diagnostic criteria for PTSD and received Cognitive Processing Therapy (CPT) in a feasibility study that preceded a subsequent randomized controlled trial. Data collection occurred between December 2012 and August 2017^4,5^.

**R-2 (Eye Movement Desensitization and Reprocessing cohort):** Participants met diagnostic criteria for PTSD and received Eye Movement Desensitization and Reprocessing therapy in routine clinical practice between January 2005 and June 2024.

**O-1 (AMT online cohort)**: Participants were recruited through Amazon Mechanical Turk (AMT). Eligibility required exposure to a traumatic event meeting DSM PTSD Criterion A based on a trauma screening checklist and IES-R scores above the predefined clinical cutoff. Assessments were completed remotely through an online survey platform between August 2021 and December 2021. The cohort consisted predominantly of non-Japanese participants.

**O-2 (Macromill online cohort):** Participants were recruited through the Macromill online survey platform in Japan as part of a large-scale online survey conducted by the National Center of Neurology and Psychiatry (NCNP). Eligibility required exposure to at least one traumatic event. Participants completed the IES-R and related questionnaires through an online survey platform. Data collection occurred between November 2016 and March 2017. Cohort details have been described previously^6^.

Participants with incomplete symptom questionnaires were excluded. For analyses of within-individual covariance structure, participants were additionally required to contribute at least three observations. Although diagnostic procedures differed across cohorts, all cohorts used the same symptom instrument (IES-R), enabling harmonized symptom-level analyses across datasets.

**Sensitivity analysis on cross-cohort loading similarity**

Sensitivity analyses evaluated the robustness of principal component similarity estimates to alternative null-model and pairing specifications. Additional analyses restricted null distributions to the top two or top five principal components rather than all available components. Cohort pairings were further analyzed separately according to variance type (temporal-temporal, between-individual -between-individual, and temporal-between-individual ).

**Sensitivity analysis on hmgPCA result**

Sensitivity analyses evaluated the robustness of hmgPCA results to alternative null-model specifications. In the primary analysis, null distributions were constructed using all principal components available for cross-cohort comparison. Additional analyses restricted component pairings in the null distribution to the top two or top five principal components. These analyses tested whether the observed reproducibility of the universal axes depended on the number of principal components included in the null model.

**Supplementary Results**

**Supplementary Table 2 | Item-wise loadings of universal symptom dimensions and trauma-event referencing properties of IES-R items.**

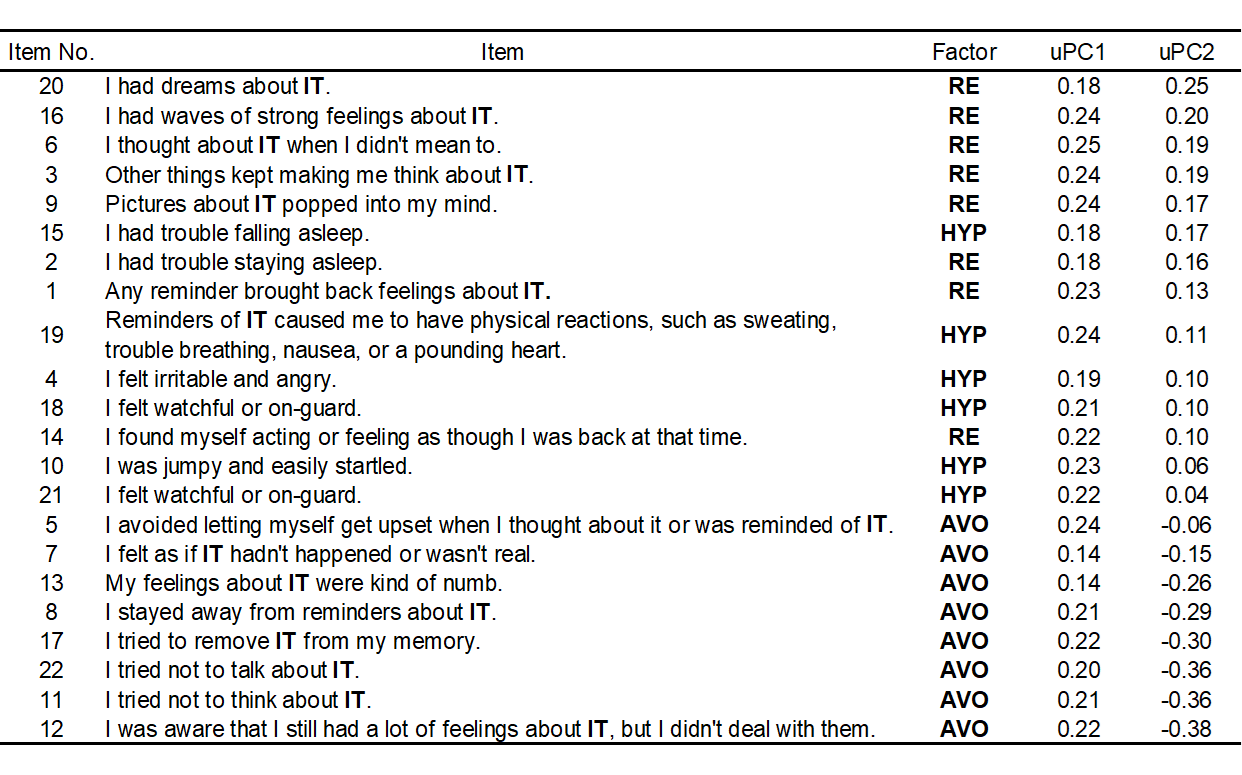

Item wording follows the original IES-R questionnaire. Items are ordered according to their loading values on uPC2, from the highest positive to the highest negative loading. “Trauma-event reference” indicates whether the item explicitly refers to the traumatic event itself (e.g., “IT”), allowing comparison of symptom items directly anchored to trauma-related experiences versus more general symptom expressions.

**Supplementary Table 3**

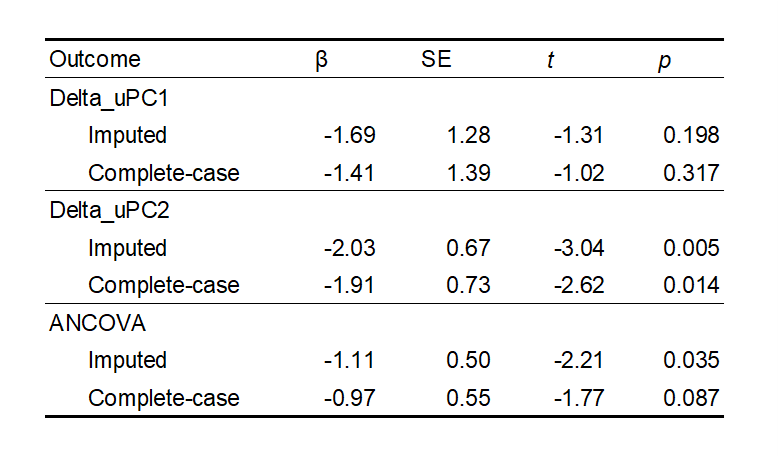

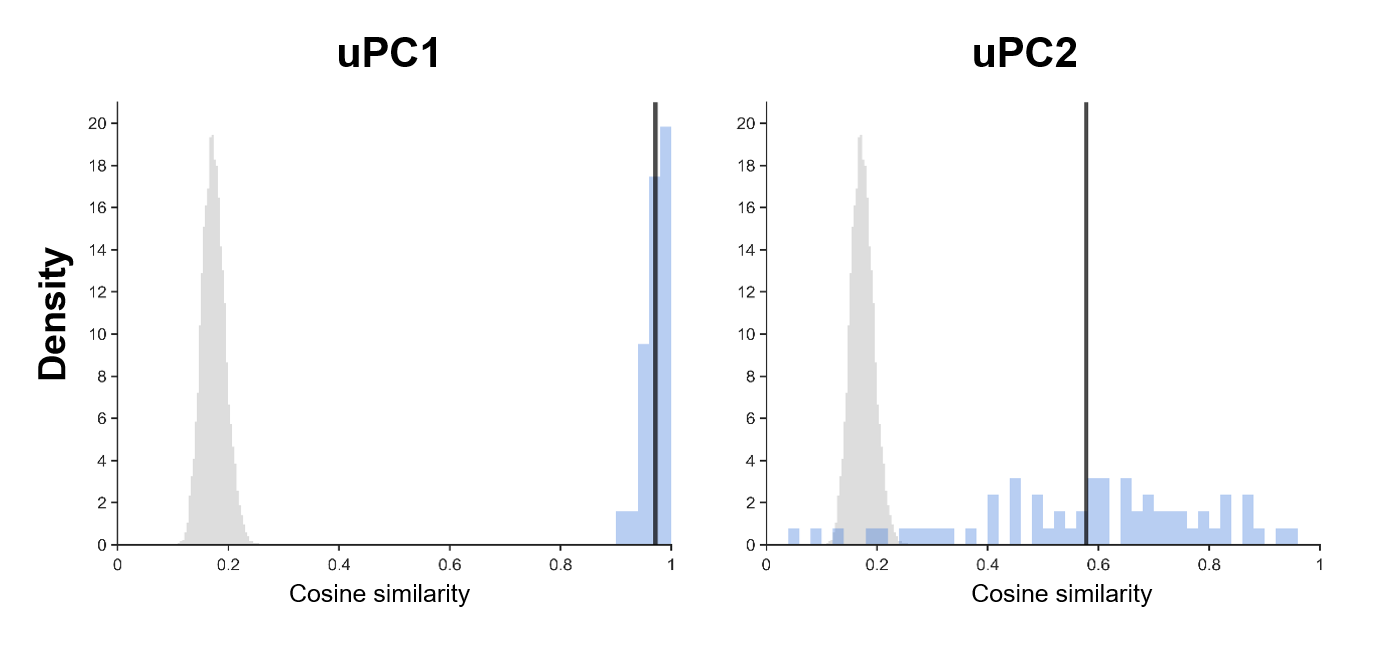

**Supplementary Fig. 1 | Similarity of within-individual and between-individual covariance across cohorts.**

Cosine similarity between loading vectors derived from within-individual covariance and those derived from between-individual covariance was evaluated across cohorts. Corresponding components showed substantially greater similarity than expected by chance, indicating that shared latent dimensions underlie both within-individual variation and between-individual heterogeneity.

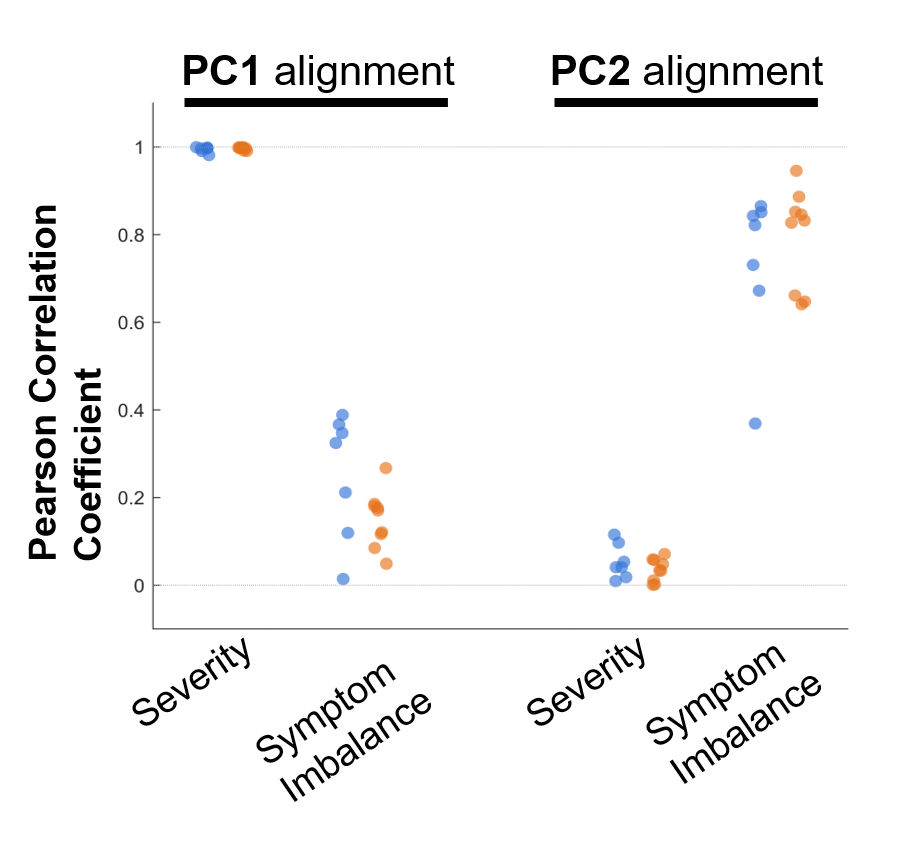

**Supplementary Fig. 2 | Robustness of external component alignment to within-individual normalization method.**

Within-individual variation was estimated using demeaned symptom trajectories instead of z-scored trajectories. PC1 remained selectively associated with overall symptom severity, whereas PC2 remained selectively associated with symptom imbalance across cohorts. Each point represents one cohort. Colors indicate variance type (blue: within-individual variation or orange: between-individual heterogeneity). Overall patterns were largely preserved relative to the primary analysis shown in **Fig. 1C**, although modest attenuation of the PC2–symptom imbalance association was observed in one cohort (R-2).

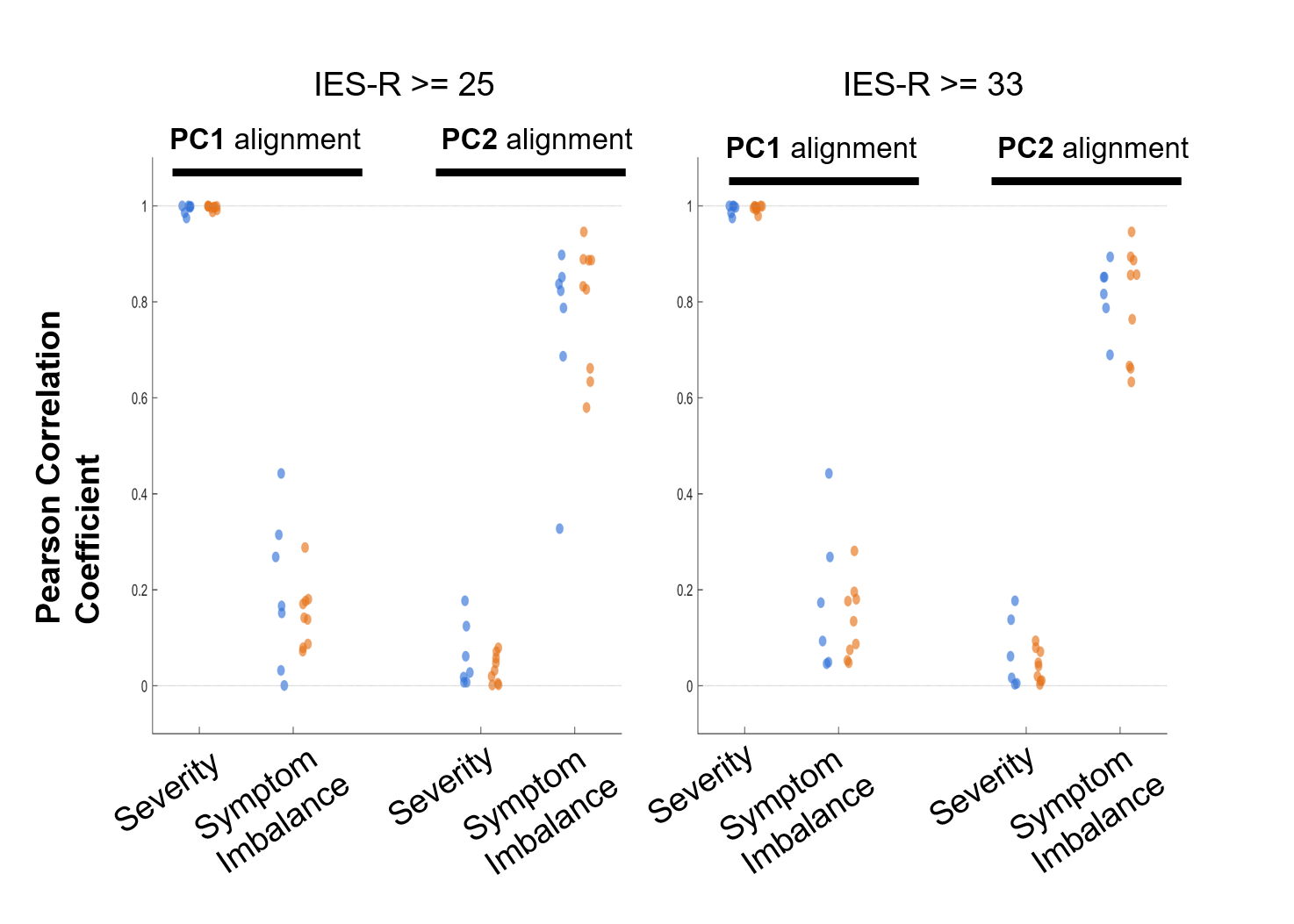

**Supplementary Fig. 3 | Robustness of the identified symptom dimensions to alternative symptom inclusion thresholds.**

Principal component analyses were repeated using more stringent symptom inclusion thresholds (IES-R ≥25 and ≥33) instead of the primary threshold (IES-R ≥20). Cohorts with fewer than 10 eligible observations after thresholding were excluded. Across thresholds, the first principal component remained strongly associated with symptom severity, whereas the second principal component remained associated with symptom imbalance. While a small number of cohorts showed increased instability due to reduced sample size, the overall loading structure and clinical interpretation were preserved, supporting the robustness of the identified low-dimensional symptom space.

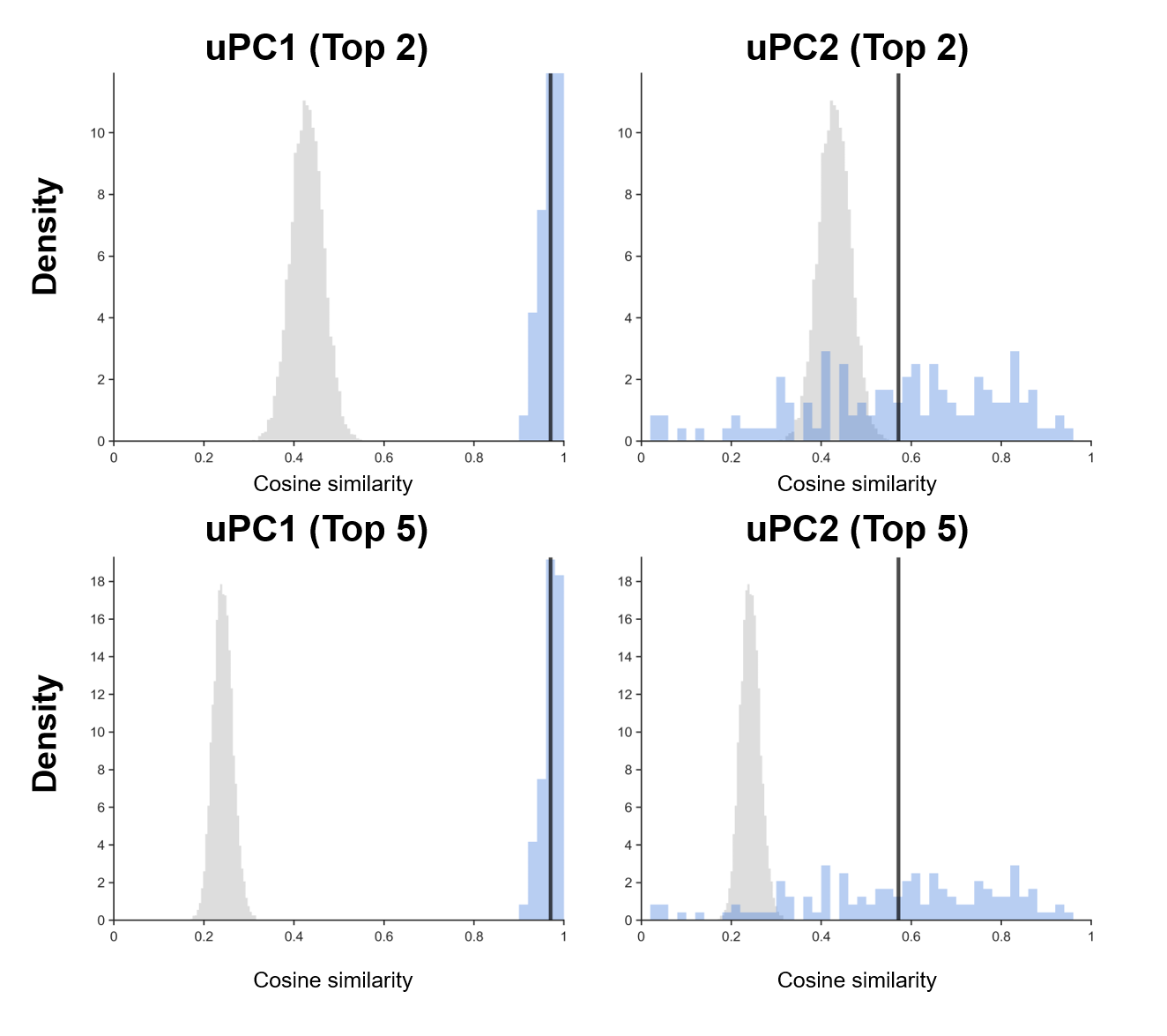

**Supplementary Fig. 4 | Robustness of cross-cohort loading similarity to null model specification.**

Principal component similarity analyses were repeated using alternative numbers of principal components. Null distributions were restricted to the top two or top five principal components rather than the full set of components included in the primary analysis. Corresponding principal components remained substantially more similar than expected by chance, indicating that the observed similarity structure was robust to the number of principal components included in the analysis.

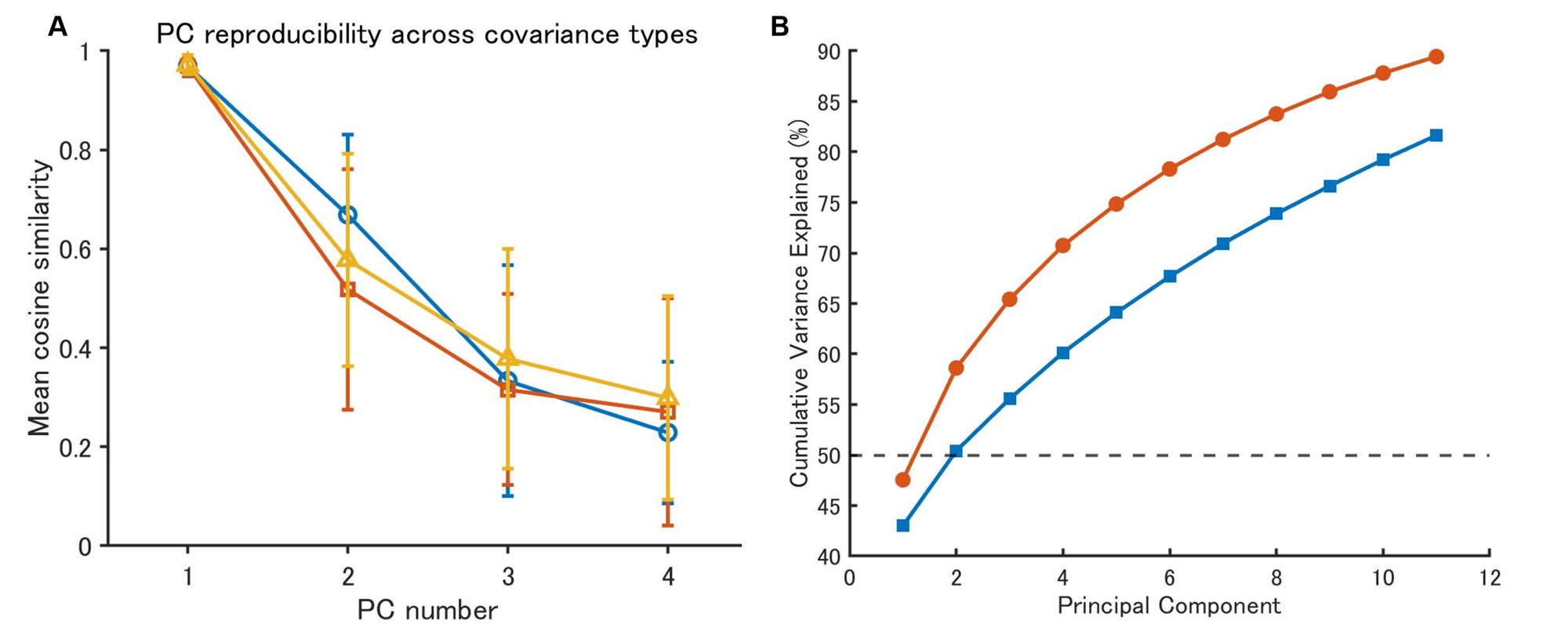

**Supplementary Fig. 5**

**| Reproducibility and dimensionality of symptom covariance structure across datasets.**

(A) Mean cross-dataset loading similarity for matched principal components (PC1–PC4), quantified using cosine similarity between loading vectors estimated independently from each covariance matrix. Blue circles indicate comparisons among temporal covariance matrices (within-individual symptom variation), orange squares indicate comparisons among between-individual covariance matrices (between-individual symptom differences), and yellow triangles indicate comparisons between temporal and between-individual covariance matrices. For each PC, cosine similarity was calculated across all possible pairs of covariance matrices within each comparison category. Error bars indicate standard deviations across pairwise comparisons.

**(B)** Mean cumulative variance explained by principal components across covariance matrices. In both between-individual covariance matrices (orange; between-individual heterogeneity) and temporal covariance matrices (blue; within-individual variation), cumulative variance explained first exceeded 50% after the second principal component, indicating that the dominant covariance structure was captured largely by the first two components. The dashed line indicates 50% cumulative variance explained.

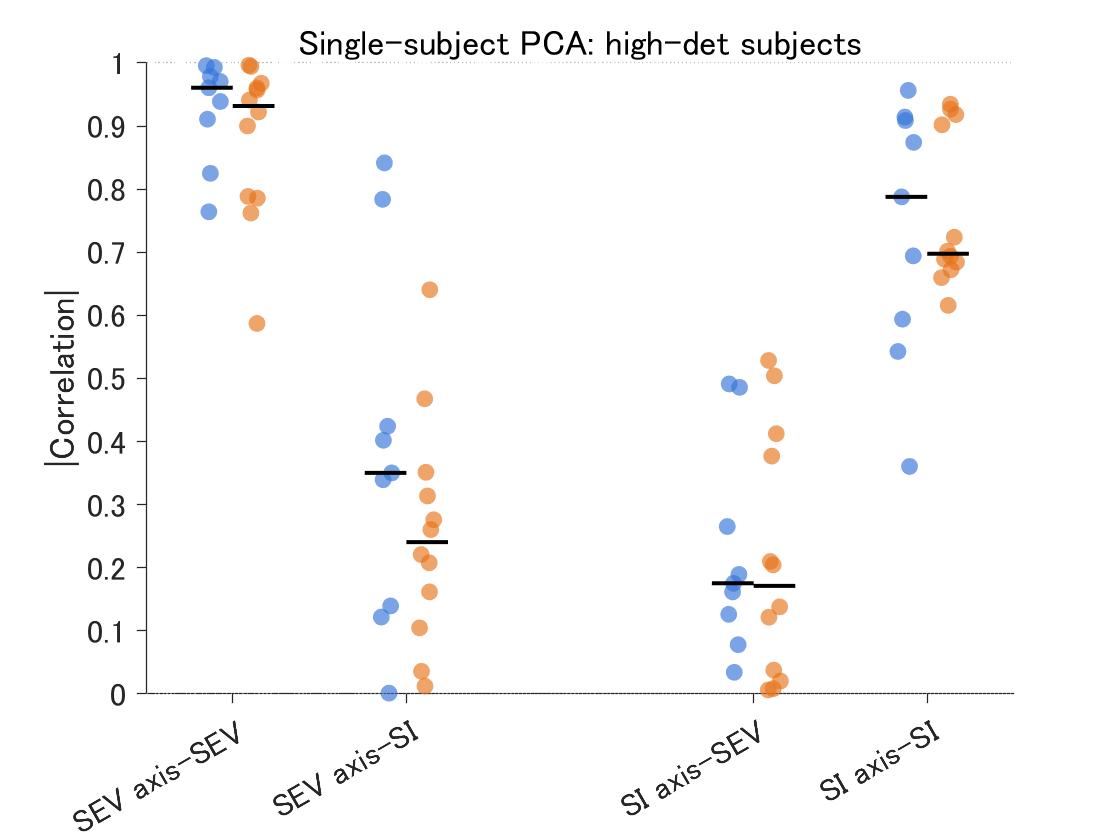

**Supplementary Fig. 6 | Single-participant recovery of severity and symptom-imbalance dimensions in ultra-dense longitudinal cohorts M-1 and M-2.**

PCA was performed separately for each participant in two independent ultra-dense longitudinal cohorts to test whether the group-level within-person covariance structure could be recovered at the individual level. Participants were included in this plot if the first two PCs showed separable correspondence with the severity and symptom-imbalance dimensions, defined as |det(R)| > 0.5, where R denotes the 2 × 2 matrix of correlations between PC1/PC2 scores and severity/symptom imbalance. PC1 and PC2 were allowed to swap order so that the component most strongly associated with severity was displayed as the SEV axis and the other as the SI axis. A total of 9/12 participants from cohort M-1 and 12/22 participants from cohort M-2 met this criterion and are shown. Dots indicate individual participants; horizontal bars indicate medians.

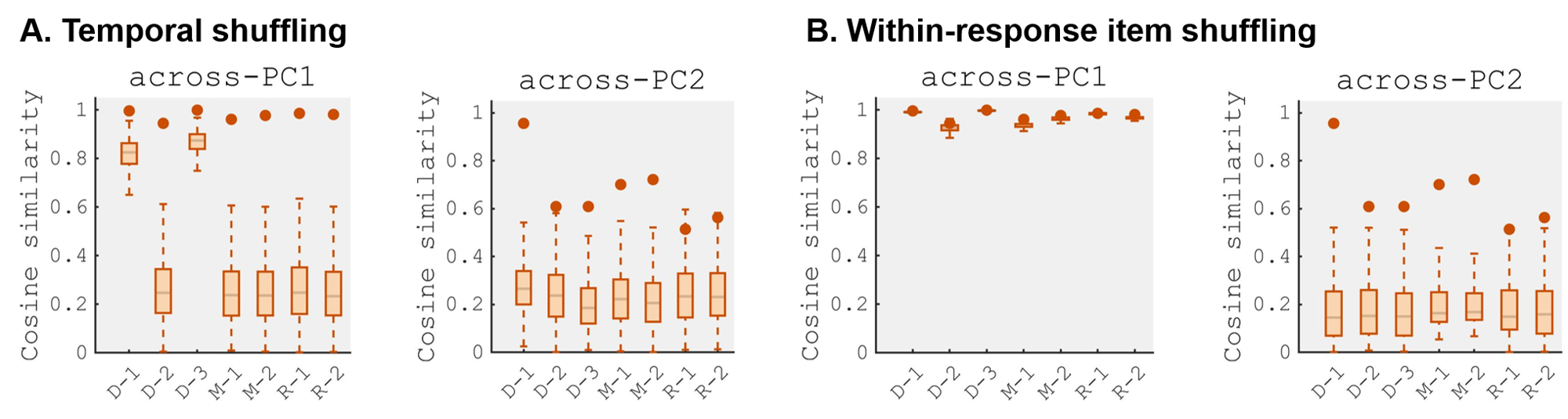

**Supplementary Fig. 7 | Shared latent symptom dimensions cannot be explained by trivial temporal or item-level statistical dependencies.**

Two complementary permutation analyses were performed to test whether the similarity between within-individual and between-individual principal component structure could arise from trivial temporal or item-level statistical dependencies (1,000 permutations per cohort). **A, Temporal shuffling:** For each participant, symptom scores for each IES-R item were independently shuffled across time, thereby disrupting temporal coordination among symptoms while preserving the empirical distribution of each symptom within that participant. Principal components were recomputed from the permuted within-individual covariance matrices, and cosine similarity with the empirically observed between-individual principal components was calculated. **B, Item identity shuffling:** Within each time-point observation, item scores were randomly shuffled across IES-R items, preserving overall response-level severity while disrupting item-specific covariance structure. Principal components were then recomputed for both within-individual and between-individual covariance matrices, and cosine similarity with the empirically observed between-individual principal components was evaluated. Dots indicate empirically observed similarity. Boxplots represent permutation-based null distributions. Across cohorts and permutation methods, observed similarity frequently exceeded permutation-based null expectations, indicating that the observed correspondence between within-individual and between-individual covariance structure cannot be explained by trivial temporal dependencies or simple response-level statistical structure alone. The main exceptions were PC1 under within-response item shuffling in cohorts D-1 (p = 0.37) and R-2 (p = 0.10), consistent with the interpretation that the first principal component primarily reflects overall symptom severity, which depends less strongly on item-specific covariance patterns because all symptom items contribute in the same direction.

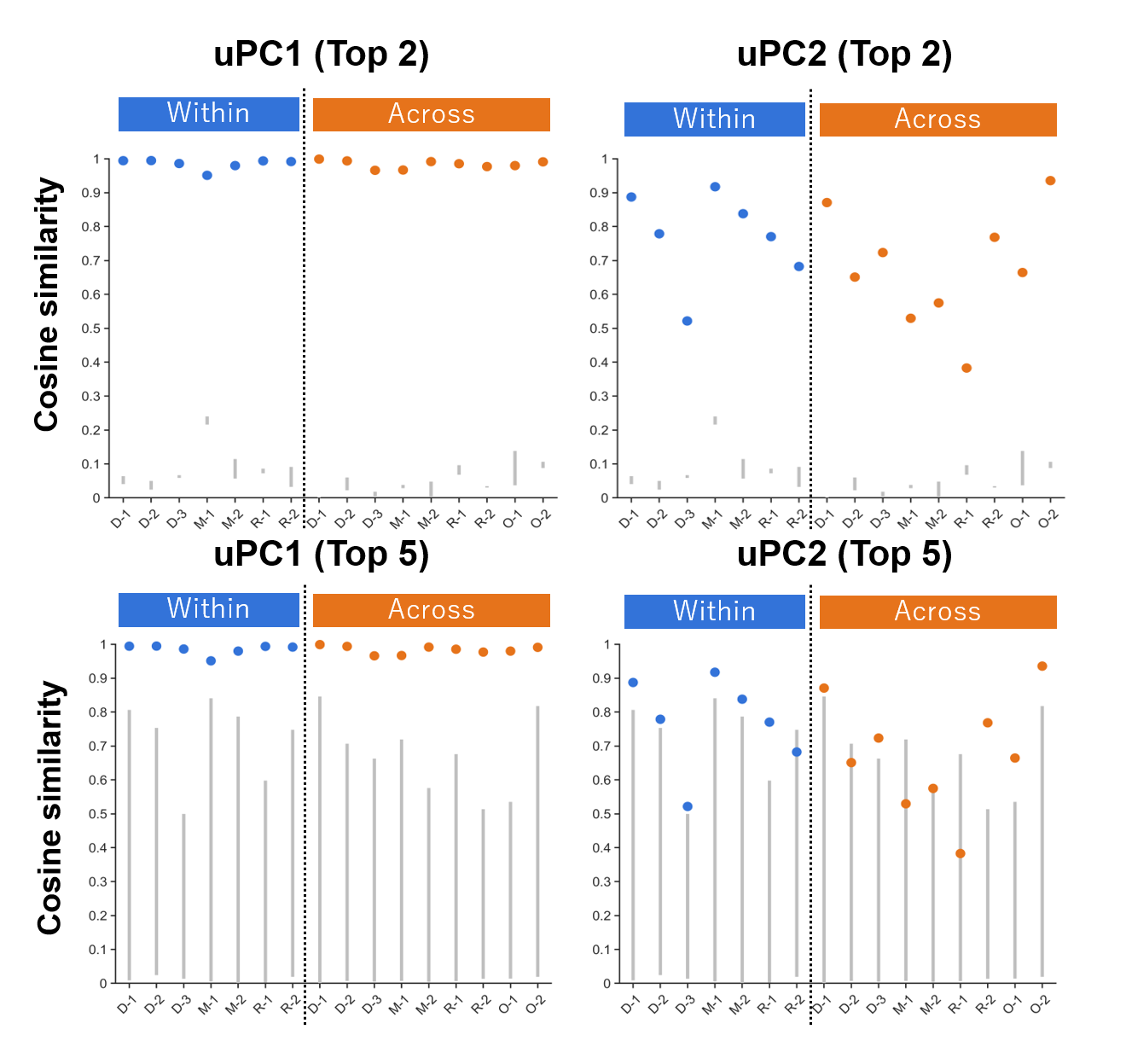

**Supplementary Fig. 8 | Cohort-wise leave-one-cohort-out generalization performance of universal axes.**

For each held-out cohort, universal loading vectors estimated from the remaining covariance matrices were compared with cohort-specific loading vectors using cosine similarity. Similarity values are shown separately for uPC1 and uPC2. Although generalization performance varied across covariance matrices, both universal axes showed consistently greater similarity than expected by chance, indicating that the shared latent dimensions were reproducible across heterogeneous covariance matrices spanning different clinical phases and sampling schemes.

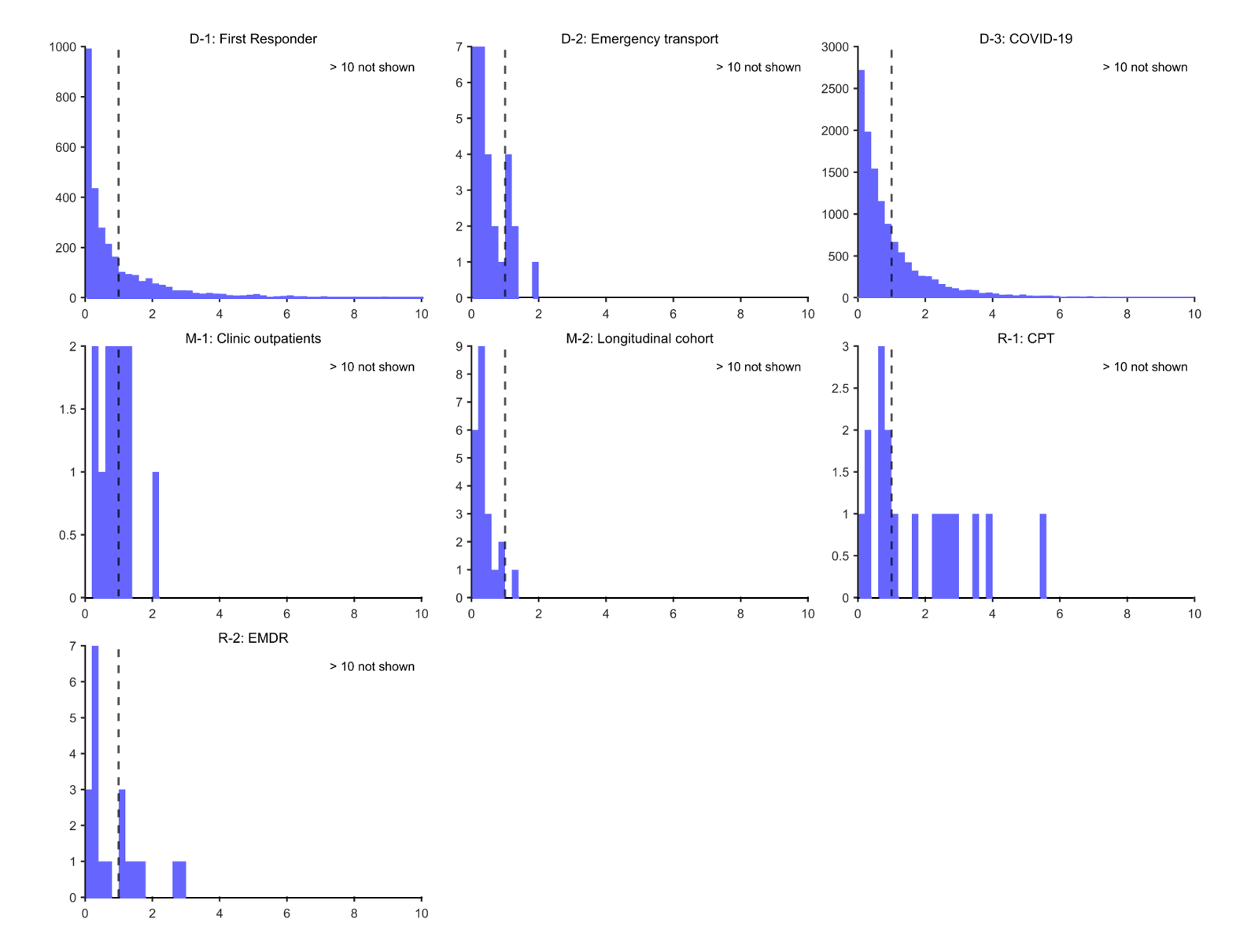

**Supplementary Fig. 9 | Distribution of within-/between-individual variance ratios across cohorts.**
 For each participant, the variance ratio was calculated as the within-individual variance along uPC2 divided by the variance of between-individual heterogeneity along uPC2 estimated at the cohort level. Distributions are shown separately for all cohorts included in the analysis. Consistent with the representative examples shown in Fig. 4B,C, considerable within-individual variation was observed across cohorts, with many participants exhibiting variance ratios greater than one, indicating that within-individual variation equaled or exceeded between-individual variances.

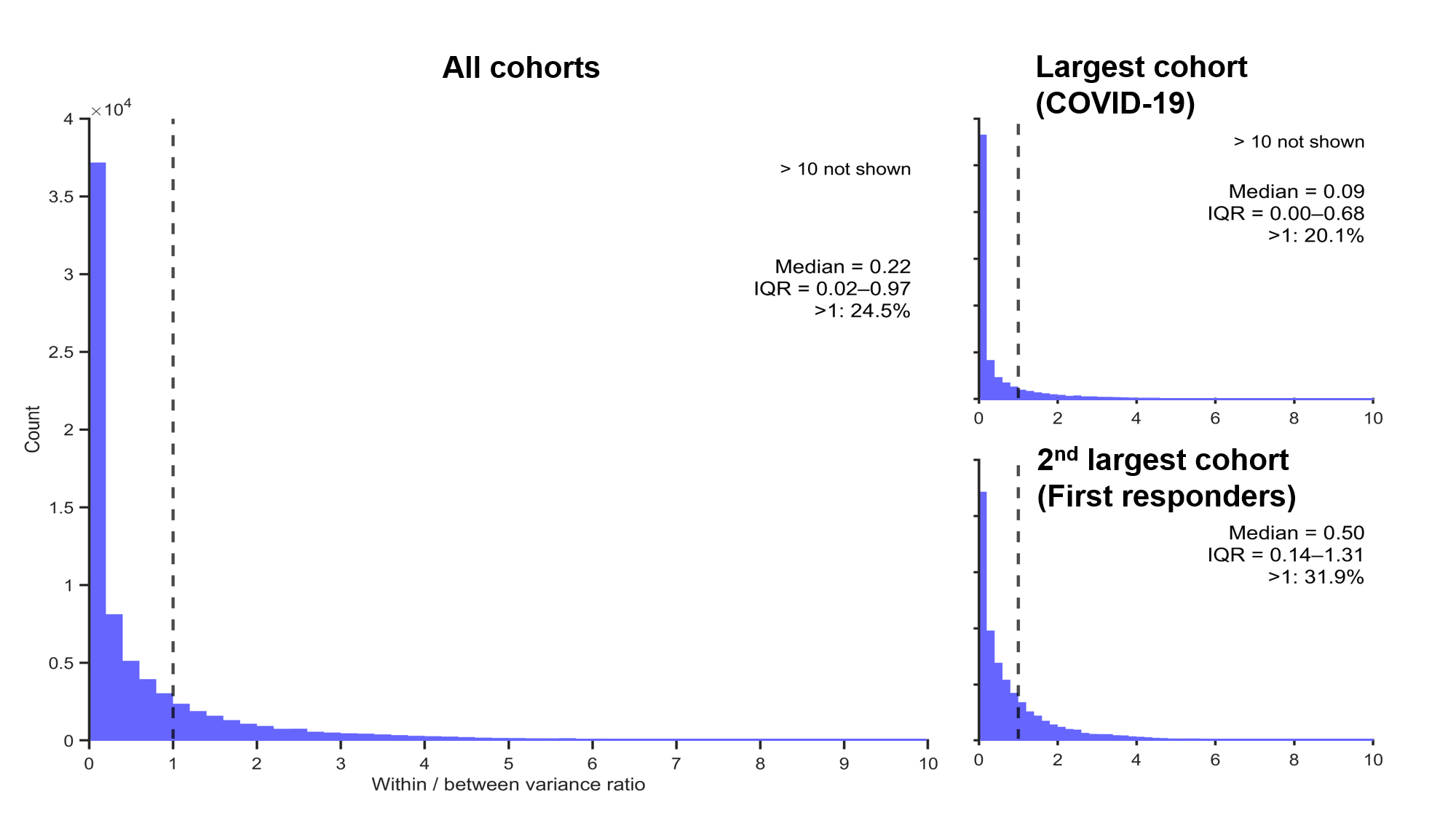

**Supplementary Fig. 10 | Robustness of variance-ratio distributions to symptom-severity inclusion criteria.**

Variance-ratio analyses were repeated using all available participants rather than restricting analyses to individuals who exceeded the clinical threshold (IES-R ≥ 20) at least once. Although the proportion of participants with variance ratios greater than one was reduced, the overall distributional pattern remained qualitatively similar, indicating that substantial within-individual variation along uPC2 was not solely driven by clinically severe cases.

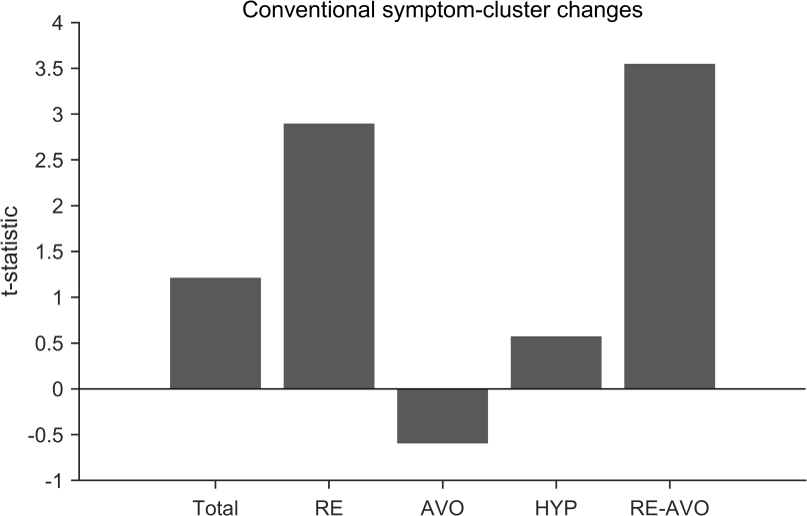

**Supplementary Fig. 11 | Comparison with conventional symptom-cluster analyses.**

To examine whether similar treatment-related differences could be detected using conventional symptom-cluster measures, we additionally compared absolute pre–post symptom changes; change by EMDR minus change by CPT. EMDR showed numerically greater overall symptom improvement, although the between-group difference did not reach significance (*t* = 1.21, *df* = 34, *p* = 0.23). Consistent with this trend, EMDR also showed significantly greater reduction in re-experiencing symptoms (*t* = 2.90, *df* = 34, *p* = 0.007). In contrast, avoidance symptoms showed a numerical trend in the opposite direction, with relatively greater reduction following CPT, although this difference was not significant (*t* = -0.60, *df* = 34, *p* = 0.56). Consequently, the predefined symptom imbalance score (ΔRE – ΔAVO) showed a larger between-group difference (*t* = 3.55, *df* = 34, *p* = 0.001) than those in re-experiencing symptoms alone, raising the possibility that CPT may preferentially improve avoidance-related symptoms relative to EMDR. Although this interpretation remains exploratory and requires prospective validation, these findings suggest that treatment-related differences may be obscured by strong severity-related effects in conventional symptom-cluster analyses, and may only become apparent after isolating relative symptom organization independently of overall symptom improvement.
